## Supplementary material for "Quantifying prevalence and risk factors of HIV multiple infection in Uganda from population-based deep-sequence data": S1 File

Supplementary File 1: Supplementary methods

Michael A. Martin[1,†], Andrea Brizzi[2], Xiaoyue Xi[2,3], Ronald Moses Galiwango[4], Sikhulile Moyo[5,6], Deogratius Ssemwanga[7,8] Alexandra Blenkinsop[2], Andrew D. Redd[9,10,11], Lucie Abeler-Dörner[12], Christophe Fraser[12], Steven J. Reynolds[4,9,10], Thomas C. Quinn[4,9,10], Joseph Kagaayi[4,13], David Bonsall[14], David Serwadda[4], Gertrude Nakigozi[4], Godfrey Kigozi[4], M. Kate Grabowski[1,4,15,†], Oliver Ratmann[2,†], with the PANGEA-HIV Consortium and the Rakai Health Sciences Program

**1** Department of Pathology, Johns Hopkins School of Medicine, Baltimore, MD, USA
**2** Department of Mathematics, Imperial College London, London, United Kingdom
**3** Medical Research Council Biostatistics Unit, University of Cambridge, Cambridge, UK
**4** Rakai Health Sciences Program, Kalisizo, Uganda
**5** Botswana Harvard AIDS Institute Partnership, Botswana Harvard HIV Reference Laboratory, Gaborone, Botswana
**6** Harvard T.H. Chan School of Public Health, Boston, MA, USA
**7** Medical Research Council/Uganda Virus Research Institute and London School of Hygiene and Tropical Medicine Uganda Research Unit, Entebbe, Uganda
**8** Uganda Virus Research Institute, Entebbe, Uganda
**9** Department of Medicine, Johns Hopkins School of Medicine, Baltimore, MD, USA
**10** Division of Intramural Research, National Institute of Allergy and Infectious Diseases, National Institutes of Health, Bethesda, MD, USA
**11** Institute of Infectious Disease and Molecular Medicine, University of Cape Town, Cape Town, South Africa
**12** Pandemic Sciences Institute, Nuffield Department of Medicine, University of Oxford, Oxford, UK
**13** Makerere University School of Public Health, Kampala, Uganda
**14** Wellcome Centre for Human Genetics, Nuffield Department of Medicine, University of Oxford, Oxford, UK
**15** Department of Epidemiology, Johns Hopkins Bloomberg School of Public Health, Baltimore, MD, USA

### 1 Supplementary methods

#### 1.1 Inference of within-host deep sequence phylogenetic trees

##### 1.1.1 Generation of putative transmission networks

To avoid running phyloscanner simultaneously on all sampled data, we first clustered participants living with HIV into putative transmission networks. Pairwise genetic distances in the form of % identity between consensus sequences (as generated by shiver) were first calculated in sliding 500 bp windows with a step size of 100 bp to account for recombination and within-host viral evolution following transmission. Pairwise genetic distances excluded sites in which either sequence had a deleted nucleotide or was unsuccessfully genotyped (e.g. an "N") or sites in which both sequences were ambiguously genotyped. Exact nucleotide matches were assigned a similarity score of 1 and partial matches based on ambiguous nucleotides in one sequence were assigned a score of 1/2 (biallelelic ambiguous nucleotide) or 1/3 (triallelic). Using genetic distance thresholds calibrated to epidemiologically confirmed transmission pairs within the RCCS [1], we clustered participants into putative transmission networks based on these pairwise distances. Inferred networks with >50 participants were decomposed into smaller networks of variable size by optimizing their modularity [2].

Finally, transmission networks were grouped into sets of sequences by 1) merging small clusters into a single sequence sets of eight participants, 2) incorporating known epidemiologically linked partners (based on RCCS survey data) for each participant in a given transmission network (regardless of genetic distance), and 3) adding 3 participants per network that were highly related to all participants in a given network on average but were not already included in the network.

##### 1.2 Base model accounting for partial sequencing success of infecting variants

To evaluate the extent to which our base model with no false-positives or false-negatives adequately described the observed data, we calculated an expected value for $\theta_i$ and consequently $(M_i^{\text{obs}}|M_i = 1)$ given an observed value for $N_i^{\text{obs}}$:

$$E[\theta_i|N_i^{\text{obs}}, M_i = 1] = 1 - \sqrt{1 - \frac{N_i^{\text{obs}}}{n^{\max}}} \quad (1)$$

$$E[M_i^{\text{obs}}|\theta_i, M_i = 1] = n^{\max}\theta_i^2. \quad (2)$$
