## Supplementary material for "Quantifying prevalence and risk factors of HIV multiple infection in Uganda from population-based deep-sequence data": S4 File

#### Supplementary File 4: Bayesian model fit diagnostics

Michael A. Martin[1,†], Andrea Brizzi[2], Xiaoyue Xi[2,3], Ronald Moses Galiwango[4], Sikhulile Moyo[5,6], Deogratius Ssemwanga[7,8] Alexandra Blenkinsop[2], Andrew D. Redd[9,10,11], Lucie Abeler-Dörner[12], Christophe Fraser[12], Steven J. Reynolds[4,9,10], Thomas C. Quinn[4,9,10], Joseph Kagaayi[4,13], David Bonsall[14], David Serwadda[4], Gertrude Nakigozi[4], Godfrey Kigozi[4], M. Kate Grabowski[1,4,15,†], Oliver Ratmann[2,†], with the PANGEA-HIV Consortium and the Rakai Health Sciences Program

**1** Department of Pathology, Johns Hopkins School of Medicine, Baltimore, MD, USA
**2** Department of Mathematics, Imperial College London, London, United Kingdom
**3** Medical Research Council Biostatistics Unit, University of Cambridge, Cambridge, UK
**4** Rakai Health Sciences Program, Kalisizo, Uganda
**5** Botswana Harvard AIDS Institute Partnership, Botswana Harvard HIV Reference Laboratory, Gaborone, Botswana
**6** Harvard T.H. Chan School of Public Health, Boston, MA, USA
**7** Medical Research Council/Uganda Virus Research Institute and London School of Hygiene and Tropical Medicine Uganda Research Unit, Entebbe, Uganda
**8** Uganda Virus Research Institute, Entebbe, Uganda
**9** Department of Medicine, Johns Hopkins School of Medicine, Baltimore, MD, USA
**10** Division of Intramural Research, National Institute of Allergy and Infectious Diseases, National Institutes of Health, Bethesda, MD, USA
**11** Institute of Infectious Disease and Molecular Medicine, University of Cape Town, Cape Town, South Africa
**12** Pandemic Sciences Institute, Nuffield Department of Medicine, University of Oxford, Oxford, UK
**13** Makerere University School of Public Health, Kampala, Uganda
**14** Wellcome Centre for Human Genetics, Nuffield Department of Medicine, University of Oxford, Oxford, UK
**15** Department of Epidemiology, Johns Hopkins Bloomberg School of Public Health, Baltimore, MD, USA

### List of Figures


1 **MCMC trace plots for parameters in base model fit to simulated data with partial sequencing success and no false positive or false negative multiple subgraph windows.** Independent chains are shown in shades of grey. Warm-up iterations are excluded. MI = multiple infection. VL = viral load ($\log_{10}$ copies/mL) standardized to mean = 0 and std. dev = 1. Std. dev. = standard deviation. . . . . . . . . . . . . . . . . . . . . . . . . . . . 6

2 **MCMC pairs plots for parameters in base model fit to simulated data with partial sequencing success and no false positive or false negative multiple subgraph windows.** Independent chains are shown in shades of grey. Warm-up iterations are excluded. Includes a sample of 250 iterations per chain. MI = multiple infection. . . . . . . . . . . . . . 7

3 **Comparison of posterior and prior distributions of parameters in base model fit to simulated data with partial sequencing success and no false positive or false negative multiple subgraph windows.** Median is plotted as the central estimate and horizontal bars extend to the 95% and 50% HPD. Some horizontal bars do not extend beyond the central point. Warm-up iterations are excluded. MI = multiple infection. VL = viral load ($\log_{10}$ copies/mL) standardized to mean = 0 and std. dev = 1. Std. dev. = standard deviation. 8

4 **MCMC trace plots for parameters in base model fit to simulated data with partial sequencing success and false positive and false negative multiple subgraph windows.** Independent chains are shown in shades of grey. Warm-up iterations are excluded. MI = multiple infection. VL = viral load ($\log_{10}$ copies/mL) standardized to mean = 0 and std. dev = 1. Std. dev. = standard deviation. . . . . . . . . . . . . . . . . . . . . . . . . . . . . . . . 8

5 **MCMC pairs plots for parameters in base model fit to simulated data with partial sequencing success and false positive and false negative multiple subgraph windows trace.** Independent chains are shown in shades of grey. Warm-up iterations are excluded. Includes a sample of 250 iterations per chain. MI = multiple infection. . . . . . . . . . . . . . 9

6 **Comparison of posterior and prior distributions of parameters in base model fit to simulated data with partial sequencing success and false positive or false negative multiple subgraph windows.** Median is plotted as the central estimate and horizontal bars extend to the 95% and 50% HPD. Some horizontal bars do not extend beyond the central point. Warm-up iterations are excluded. MI = multiple infection. VL = viral load ($\log_{10}$ copies/mL) standardized to mean = 0 and std. dev = 1. Std. dev. = standard deviation. . . 10

7 **MCMC trace plots for parameters in full model fit to simulated data with partial sequencing success and false positive and false negative multiple subgraph windows.** Independent chains are shown in shades of grey. Warm-up iterations are excluded.MI = multiple infection. VL = viral load ($\log_{10}$ copies/mL) standardized to mean = 0 and std. dev = 1. Std. dev. = standard deviation. . . . . . . . . . . . . . . . . . . . . . . . . . . . . . . . . . . 10

8 **MCMC pairs plots for parameters in full model fit to simulated data with partial sequencing success and false positive and false negative multiple subgraph windows trace.** Independent chains are shown in shades of grey. Warm-up iterations are excluded. MI = multiple infection. . . . . . . . . . . . . . . . . . . . . . . . . . . . . . . . . . . . . . 11

9 **Comparison of posterior and prior distributions of parameters in full model fit to simulated data with partial sequencing success and false positive or false negative multiple subgraph windows.** Median is plotted as the central estimate and horizontal bars extend to the 95% and 50% HPD. Some horizontal bars do not extend beyond the central point. Warm-up iterations are excluded. MI = multiple infection. VL = viral load ($\log_{10}$ copies/mL) standardized to mean = 0 and std. dev = 1. Std. dev. = standard deviation. . . 12

10 **MCMC trace plots for parameters in extended model fit to simulated data with partial sequencing success, false positive and false negative multiple subgraph windows, and a binary risk factor for harboring multiple infection.** Independent chains are shown in shades of grey. Warm-up iterations are excluded. MI = multiple infection. VL = viral load ($\log_{10}$ copies/mL) standardized to mean = 0 and std. dev = 1. Std. dev. = standard deviation. . . . . . . . . . . . . . . . . . . . . . . . . . . . . . . . . . . . . . 13

11 **MCMC pairs plots for parameters in extended model fit to simulated data with partial sequencing success, false positive and false negative multiple subgraph windows, and a binary risk factor for harboring multiple infection.** Independent chains are shown in shades of grey. Warm-up iterations are excluded. Includes a sample of 250 iterations per chain. MI = multiple infection. . . . . . . . . . 14

12 **Comparison of posterior and prior distributions of parameters in full model fit to simulated data with partial sequencing success, false positive or false negative multiple subgraph windows, and a binary risk factor for harboring multiple infection.** Median is plotted as the central estimate and horizontal bars extend to the 95% and 50% HPD. Some horizontal bars do not extend beyond the central point. Warm-up iterations are excluded. MI = multiple infection. VL = viral load ($\log_{10}$ copies/mL) standardized to mean = 0 and std. dev = 1. Std. dev. = standard deviation. . . . . . . . . 15

13 **MCMC trace plots for parameters in full model fit to deep sequence data generated from 2,029 Rakai Community Cohort Study participants living with viremic HIV.** Independent chains are shown in shades of grey. Warm-up iterations are excluded. MI = multiple infection. VL = viral load ($\log_{10}$ copies/mL) standardized to mean = 0 and std. dev = 1. Std. dev. = standard deviation. . . . . . . . . . 16

14 **MCMC pairs plots for parameters in full model fit to deep sequence data generated from 2,029 Rakai Community Cohort Study participants living with viremic HIV.** Independent chains are shown in shades of grey. Warm-up iterations are excluded. Includes a sample of 250 iterations per chain. MI = multiple infection. . . . . . . . . . 17

15 **Comparison of posterior and prior distributions of parameters in full model fit to deep sequence data generated from 2,029 Rakai Community Cohort Study participants living with viremic HIV.** Median is plotted as the central estimate and horizontal bars extend to the 95% and 50% HPD. Some horizontal bars do not extend beyond the central point. Warm-up iterations are excluded. MI = multiple infection. VL = viral load ($\log_{10}$ copies/mL) standardized to mean = 0 and std. dev = 1. Std. dev. = standard deviation. 18

16 **MCMC trace plots for parameters in extended model with community type as a risk factor fit to deep sequence data generated from 2,029 Rakai Community Cohort Study participants living with viremic HIV.** Independent chains are shown in shades of grey. Warm-up iterations are excluded. MI = multiple infection. VL = viral load ($\log_{10}$ copies/mL) standardized to mean = 0 and std. dev = 1. Std. dev. = standard deviation. 19

17 **MCMC pairs plots for parameters in extended model with community type as a risk factor fit to deep sequence data generated from 2,029 Rakai Community Cohort Study participants living with viremic HIV.** Independent chains are shown in shades of grey. Warm-up iterations are excluded. Includes a sample of 250 iterations per chain. MI = multiple infection. . . . . . . . . . 20

18 **Comparison of posterior and prior distributions of parameters in extended model with community type as a risk factor fit to deep sequence data generated from 2,029 Rakai Community Cohort Study participants living with viremic HIV.** Median is plotted as the central estimate and horizontal bars extend to the 95% and 50% HPD. Some horizontal bars do not extend beyond the central point. Warm-up iterations are excluded. MI = multiple infection. VL = viral load ($\log_{10}$ copies/mL) standardized to mean = 0 and std. dev = 1. Std. dev. = standard deviation. . . . . . . . . . 21

19 **MCMC trace plots for parameters in extended model with deep-sequencing protocol as a risk factor fit to deep sequence data generated from 2,029 Rakai Community Cohort Study participants living with viremic HIV.** Independent chains are shown in shades of grey. Warm-up iterations are excluded. MI = multiple infection. VL = viral load ($\log_{10}$ copies/mL) standardized to mean = 0 and std. dev = 1. Std. dev. = standard deviation. . . . . . . . . . 22

20 **MCMC pairs plots for parameters in extended model withdeep-sequencing protocol as a risk factor fit to deep sequence data generated from 2,029 Rakai Community Cohort Study participants living with viremic HIV.** Independent chains are shown in shades of grey. Warm-up iterations are excluded. Includes a sample of 250 iterations per chain. MI = multiple infection. . . . . . . . . . . . . . . . . . . . . . . . . . . . . . . . . . . . . . . . 23

21 **Comparison of posterior and prior distributions of parameters in extended model with sequencing technology as a risk factor fit to deep sequence data generated from 2,029 Rakai Community Cohort Study participants living with viremic HIV.** Median is plotted as the central estimate and horizontal bars extend to the 95% and 50% HPD. Some horizontal bars do not extend beyond the central point. Warm-up iterations are excluded. MI = multiple infection. VL = viral load ($\log_{10}$ copies/mL) standardized to mean = 0 and std. dev = 1. Std. dev. = standard deviation. . . . . . . . . . . . . . . . . . . . . . . 24

22 **MCMC trace plots for parameters in extended model with deep-sequencing protocol and community type as a risk factor fit to deep sequence data generated from 2,029 Rakai Community Cohort Study participants living with viremic HIV.** Independent chains are shown in shades of grey. Warm-up iterations are excluded. MI = multiple infection. VL = viral load ($\log_{10}$ copies/mL) standardized to mean = 0 and std. dev = 1. Std. dev. = standard deviation. . . . . . . . . . . . . . . . . . . . . . . . . . . . . . 25

23 **MCMC pairs plots for parameters in extended model withdeep-sequencing protocol and community type as a risk factor fit to deep sequence data generated from 2,029 Rakai Community Cohort Study participants living with viremic HIV.** Independent chains are shown in shades of grey. Warm-up iterations are excluded. Includes a sample of 250 iterations per chain. MI = multiple infection. . . . . . . . . . . . . . . . . . . . . . . . . . . 26

24 **Comparison of posterior and prior distributions of parameters in extended model with sequencing technology and community type as a risk factor fit to deep sequence data generated from 2,029 Rakai Community Cohort Study participants living with viremic HIV.** Median is plotted as the central estimate and horizontal bars extend to the 95% and 50% HPD. Some horizontal bars do not extend beyond the central point. Warm-up iterations are excluded. MI = multiple infection. VL = viral load ($\log_{10}$ copies/mL) standardized to mean = 0 and std. dev = 1. Std. dev. = standard deviation. . . 27

25 **MCMC trace plots for parameters in extended model with community type and lifetime sex partners as a risk factor for harboring multiple infections fit to deep sequence data generated from 997 men living with viremic HIV who participated in the Rakai Community Cohort Study.** Independent chains are shown in shades of grey. Warm-up iterations are excluded. MI = multiple infection. VL = viral load ($\log_{10}$ copies/mL) standardized to mean = 0 and std. dev = 1. Std. dev. = standard deviation. . . . . . . . . . 28

26 **MCMC pairs plots for parameters in extended model with community type and lifetime sex partners as a risk factor for harboring multiple infections fit to deep sequence data generated from 997 men living with viremic HIV who participated in the Rakai Community Cohort Study.** Independent chains are shown in shades of grey. Warm-up iterations are excluded. Includes a sample of 250 iterations per chain. MI = multiple infection. . . . . . . . . . . . . . . . . . . . . . . . . . . . . . . . . . . . . . . . . . . . . . 29

27 **Comparison of posterior and prior distributions of parameters in extended model with sequencing technology as a risk factor fit to deep sequence data generated from 997 Rakai Community Cohort Study participants living with viremic HIV.** Median is plotted as the central estimate and horizontal bars extend to the 95% and 50% HPD. Some horizontal bars do not extend beyond the central point. Warm-up iterations are excluded. MI = multiple infection. VL = viral load ($\log_{10}$ copies/mL) standardized to mean = 0 and std. dev = 1. Std. dev. = standard deviation. . . . . . . . . . . . . . . . . . . . . . . 30

28 **MCMC trace plots for parameters in extended model with variable selection to identify risk factor for harboring multiple infections fit to deep sequence data generated from 1,970 Rakai Community Cohort Study participants living with viremic HIV.** Independent chains are shown in shades of grey. Warm-up iterations are excluded. MI = multiple infection. VL = viral load ($\log_{10}$ copies/mL) standardized to mean = 0 and std. dev = 1. Std. dev. = standard deviation. . . . . . . . . . . . . . . . . . . . . . . 31
29 **MCMC pairs plots for parameters in extended model with variable selection to identify risk factor for harboring multiple infections fit to deep sequence data generated from 1,970 Rakai Community Cohort Study participants living with viremic HIV.** Independent chains are shown in shades of grey. Warm-up iterations are excluded. Includes a sample of 250 iterations per chain. MI = multiple infection. . . . . . . 32
30 **Comparison of posterior and prior distributions of parameters in extended model with variable selection to identify risk factor fit to deep sequence data generated from 1,970 Rakai Community Cohort Study participants living with viremic HIV.** Median is plotted as the central estimate and horizontal bars extend to the 95% and 50% HPD. Some horizontal bars do not extend beyond the central point. Warm-up iterations are excluded. MI = multiple infection. VL = viral load ($\log_{10}$ copies/mL) standardized to mean = 0 and std. dev = 1. Std. dev. = standard deviation. . . . . . . . . . . . . . . . . . . . . . . 33

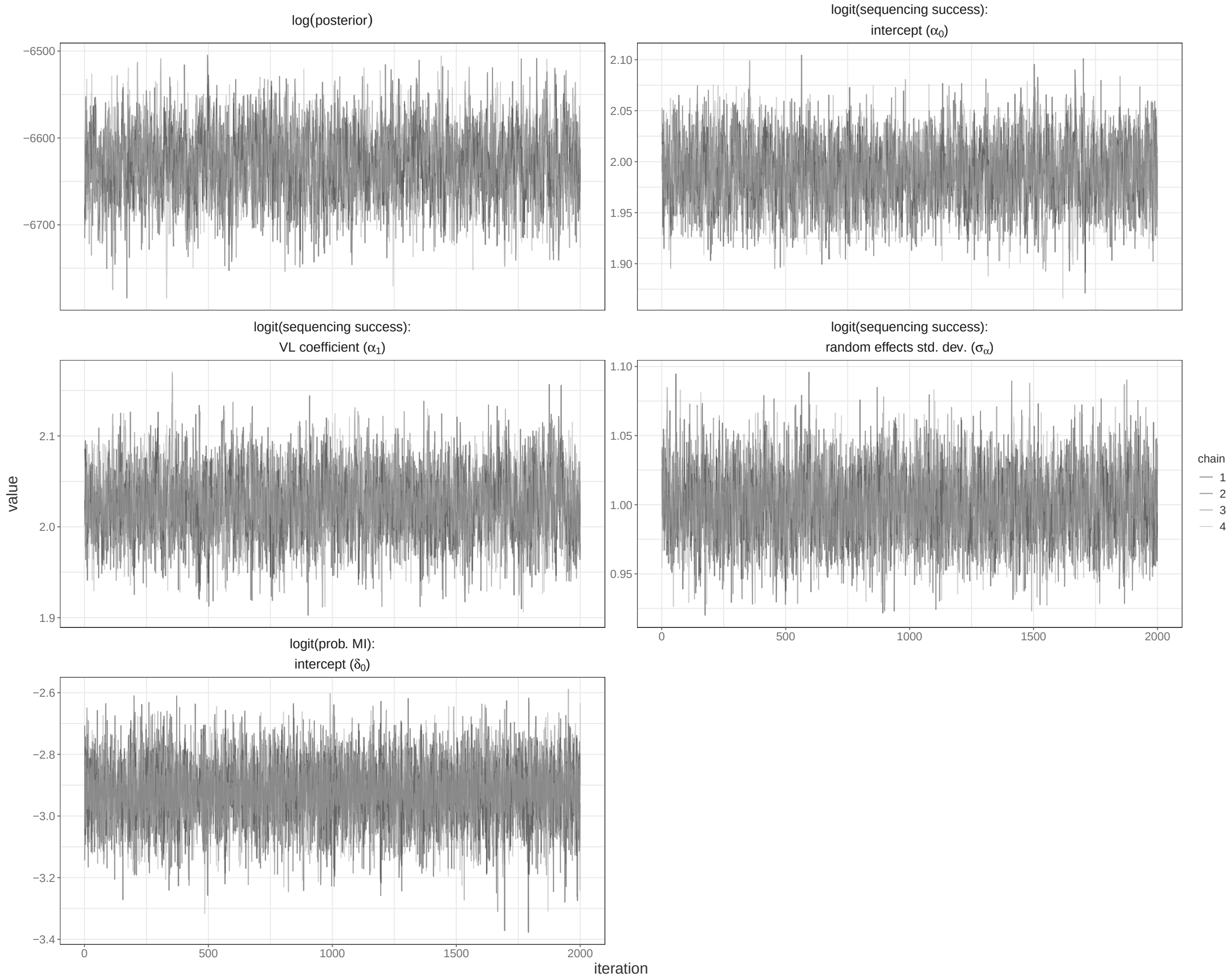


**Fig S.D. 1. MCMC trace plots for parameters in base model fit to simulated data with partial sequencing success and no false positive or false negative multiple subgraph windows.** Independent chains are shown in shades of grey. Warm-up iterations are excluded. MI = multiple infection. VL = viral load ($\log_{10}$ copies/mL) standardized to mean = 0 and std. dev = 1. Std. dev. = standard deviation.

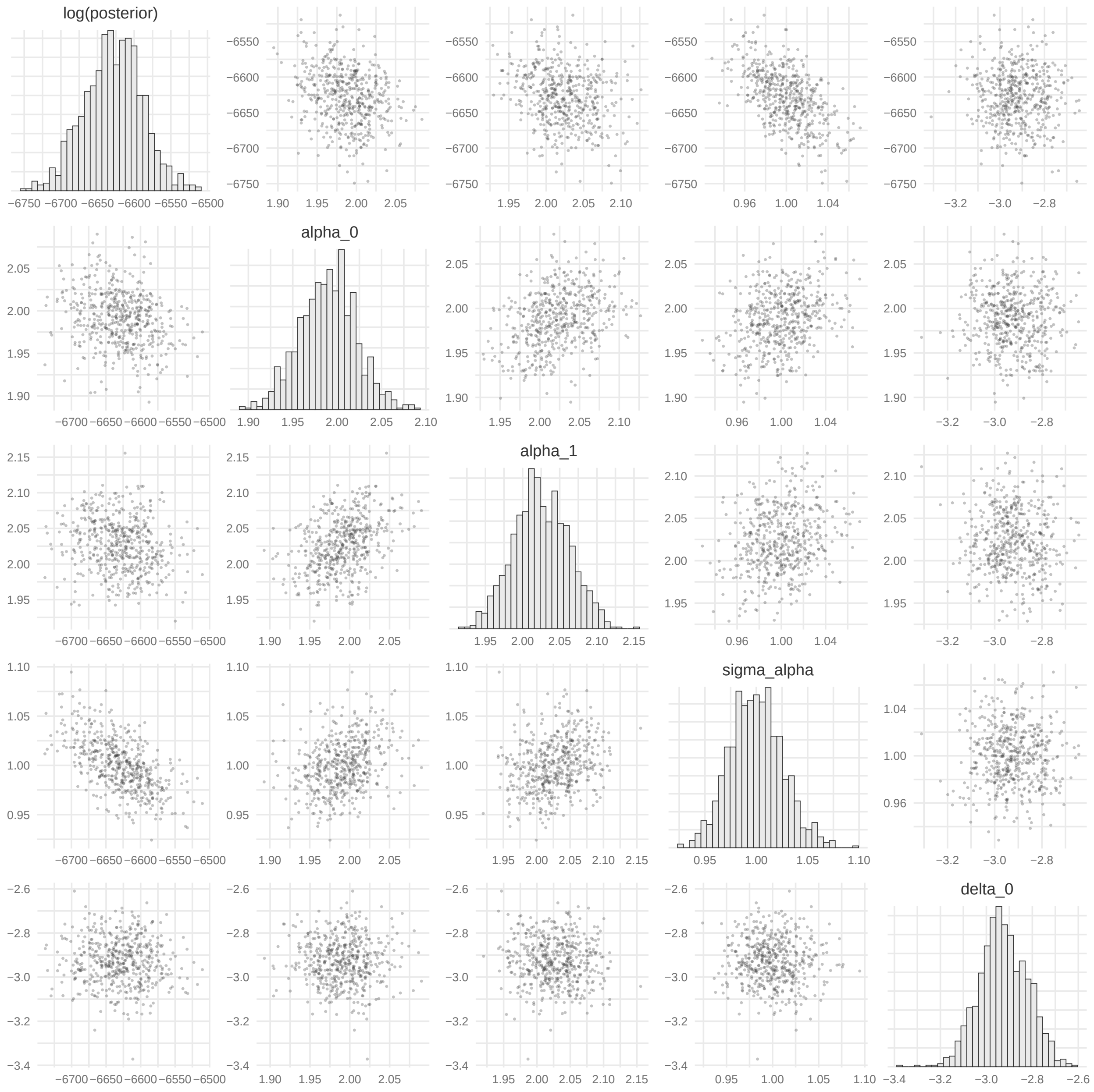


**Fig S.D. 2. MCMC pairs plots for parameters in base model fit to simulated data with partial sequencing success and no false positive or false negative multiple subgraph windows.** Independent chains are shown in shades of grey. Warm-up iterations are excluded. Includes a sample of 250 iterations per chain. MI = multiple infection.

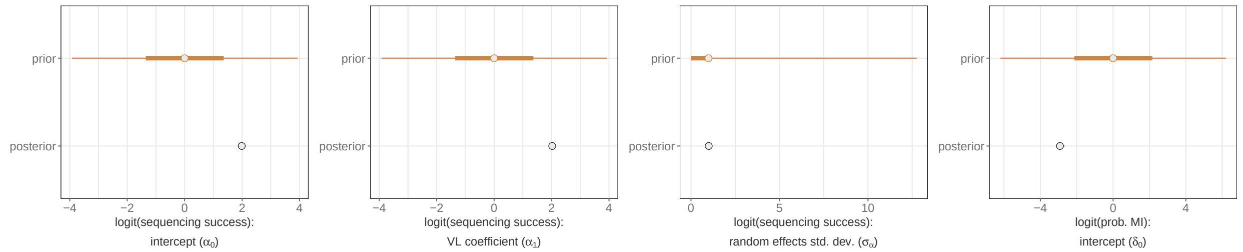


**Fig S.D. 3. Comparison of posterior and prior distributions of parameters in base model fit to simulated data with partial sequencing success and no false positive or false negative multiple subgraph windows.** Median is plotted as the central estimate and horizontal bars extend to the 95% and 50% HPD. Some horizontal bars do not extend beyond the central point. Warm-up iterations are excluded. MI = multiple infection. VL = viral load ($\log_{10}$ copies/mL) standardized to mean = 0 and std. dev = 1. Std. dev. = standard deviation.

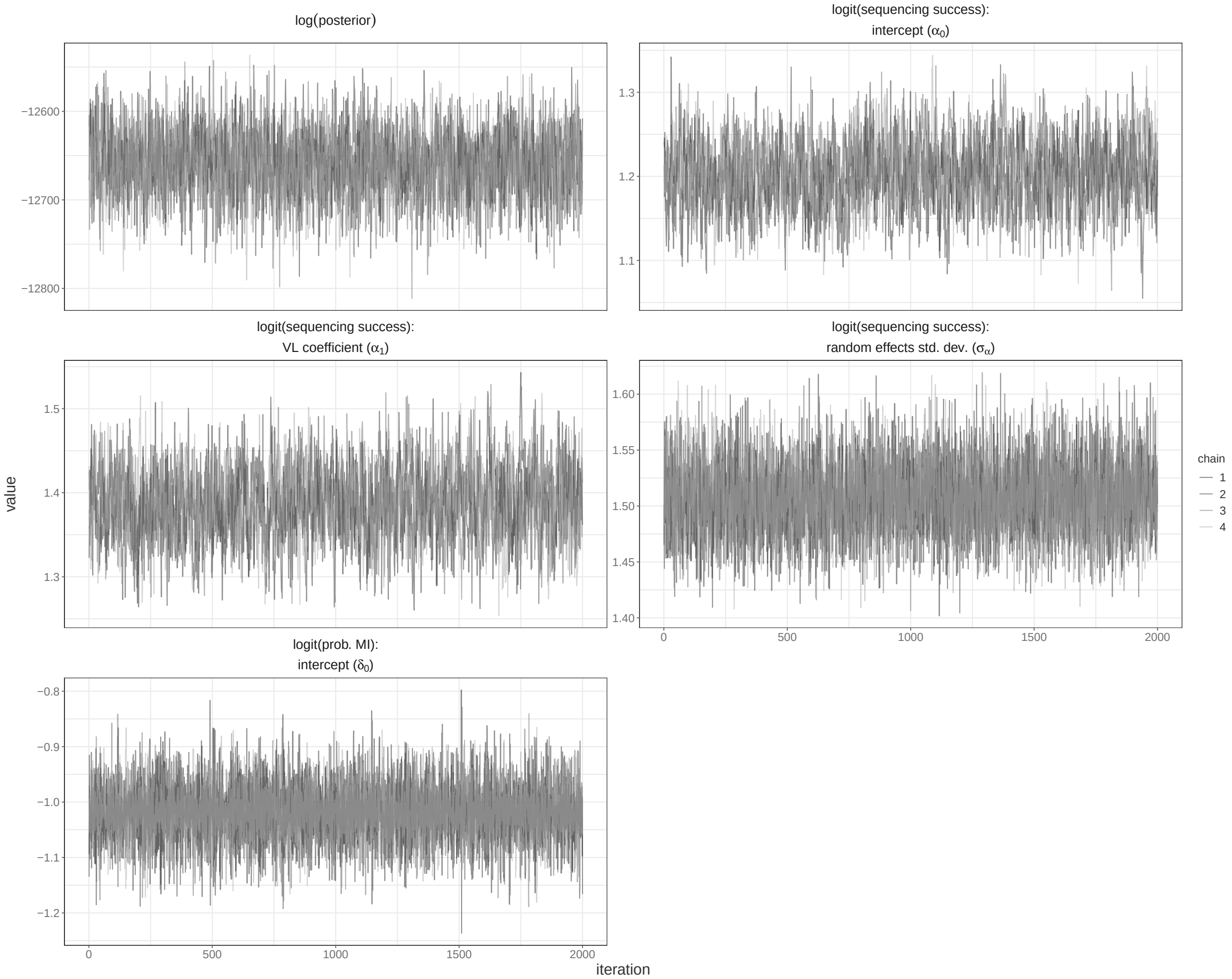


**Fig S.D. 4. MCMC trace plots for parameters in base model fit to simulated data with partial sequencing success and false positive and false negative multiple subgraph windows.** Independent chains are shown in shades of grey. Warm-up iterations are excluded. MI = multiple infection. VL = viral load ($\log_{10}$ copies/mL) standardized to mean = 0 and std. dev = 1. Std. dev. = standard deviation.

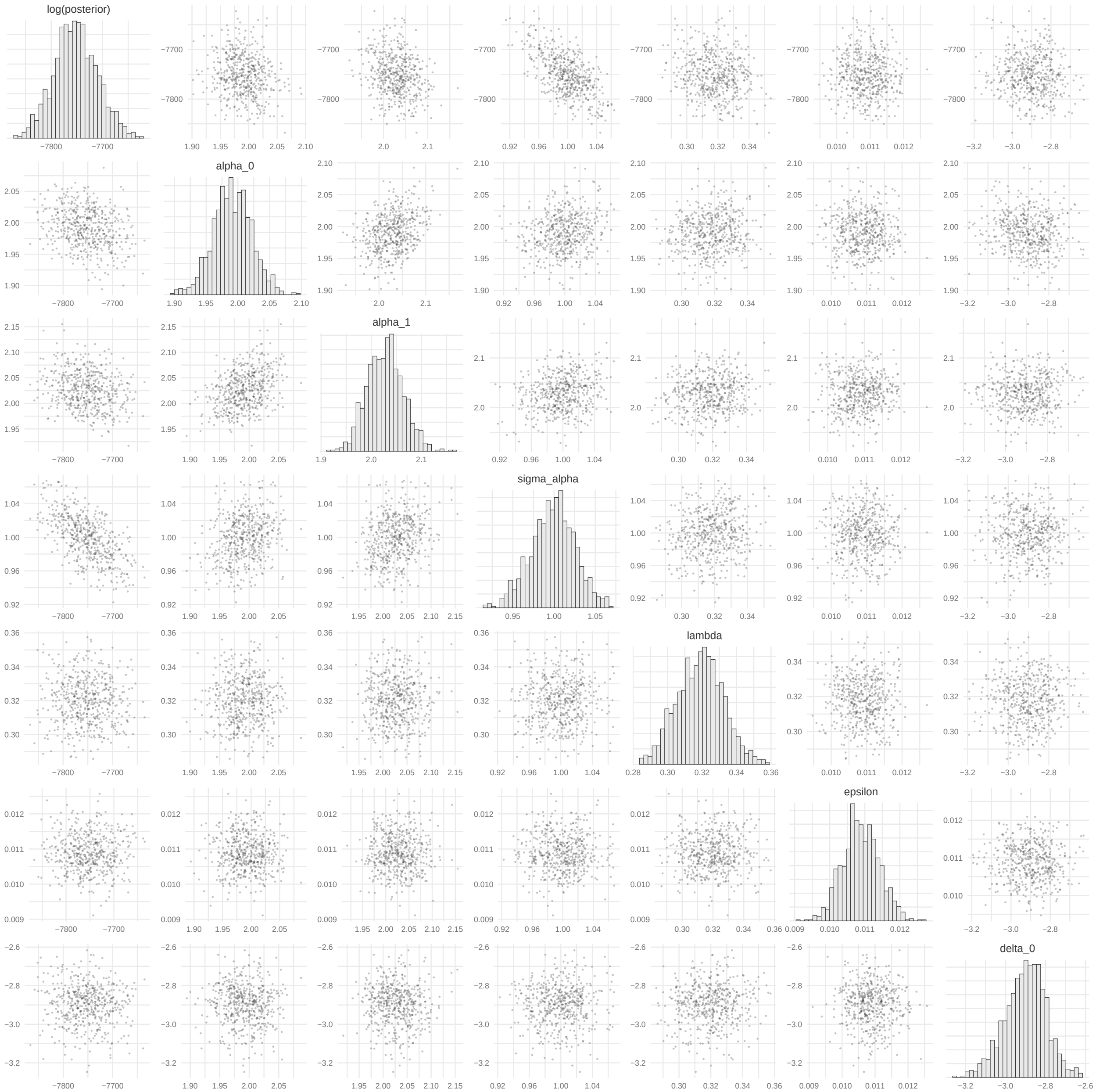


**Fig S.D. 5. MCMC pairs plots for parameters in base model fit to simulated data with partial sequencing success and false positive and false negative multiple subgraph windows trace.** Independent chains are shown in shades of grey. Warm-up iterations are excluded. Includes a sample of 250 iterations per chain. MI = multiple infection.

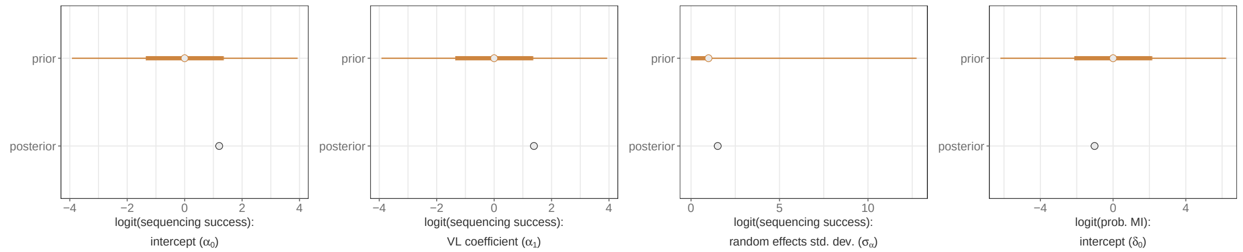


**Fig S.D. 6. Comparison of posterior and prior distributions of parameters in base model fit to simulated data with partial sequencing success and false positive or false negative multiple subgraph windows.** Median is plotted as the central estimate and horizontal bars extend to the 95% and 50% HPD. Some horizontal bars do not extend beyond the central point. Warm-up iterations are excluded. MI = multiple infection. VL = viral load ($\log_{10}$ copies/mL) standardized to mean = 0 and std. dev = 1. Std. dev. = standard deviation.

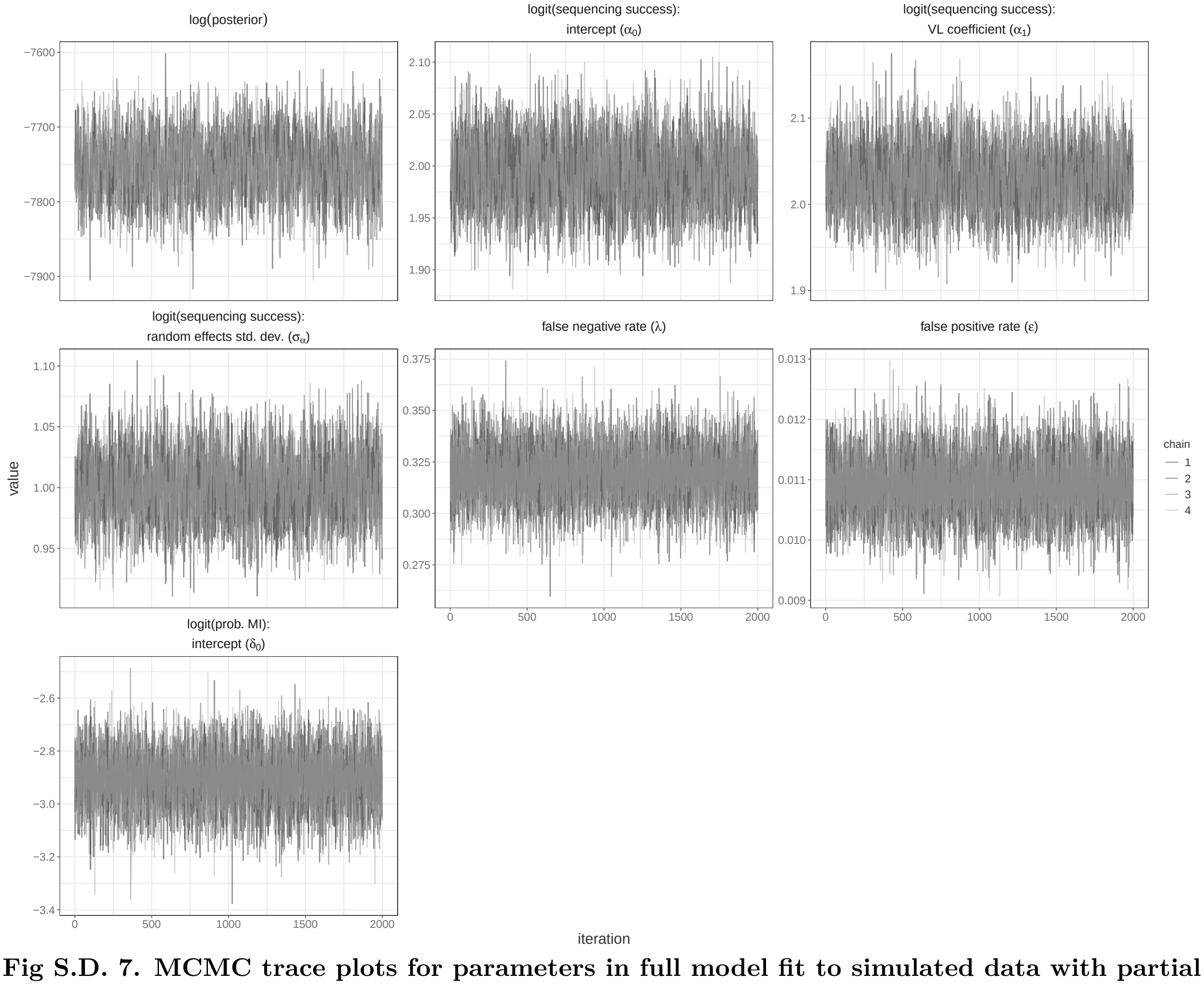


**Fig S.D. 7. MCMC trace plots for parameters in full model fit to simulated data with partial sequencing success and false positive and false negative multiple subgraph windows.** Independent chains are shown in shades of grey. Warm-up iterations are excluded.MI = multiple infection. VL = viral load ($\log_{10}$ copies/mL) standardized to mean = 0 and std. dev = 1. Std. dev. = standard deviation.

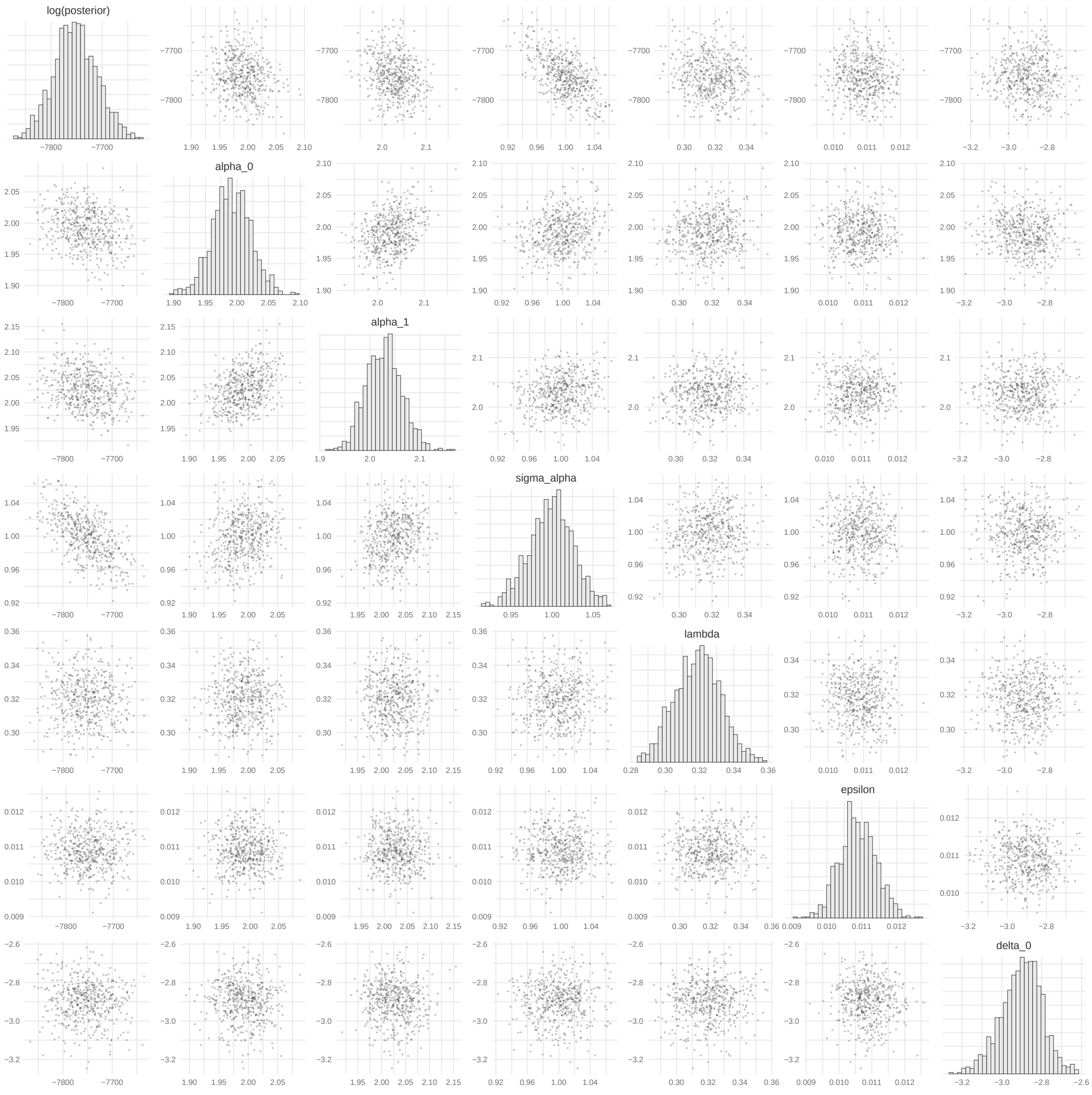


**Fig S.D. 8. MCMC pairs plots for parameters in full model fit to simulated data with partial sequencing success and false positive and false negative multiple subgraph windows trace.** Independent chains are shown in shades of grey. Warm-up iterations are excluded. MI = multiple infection.

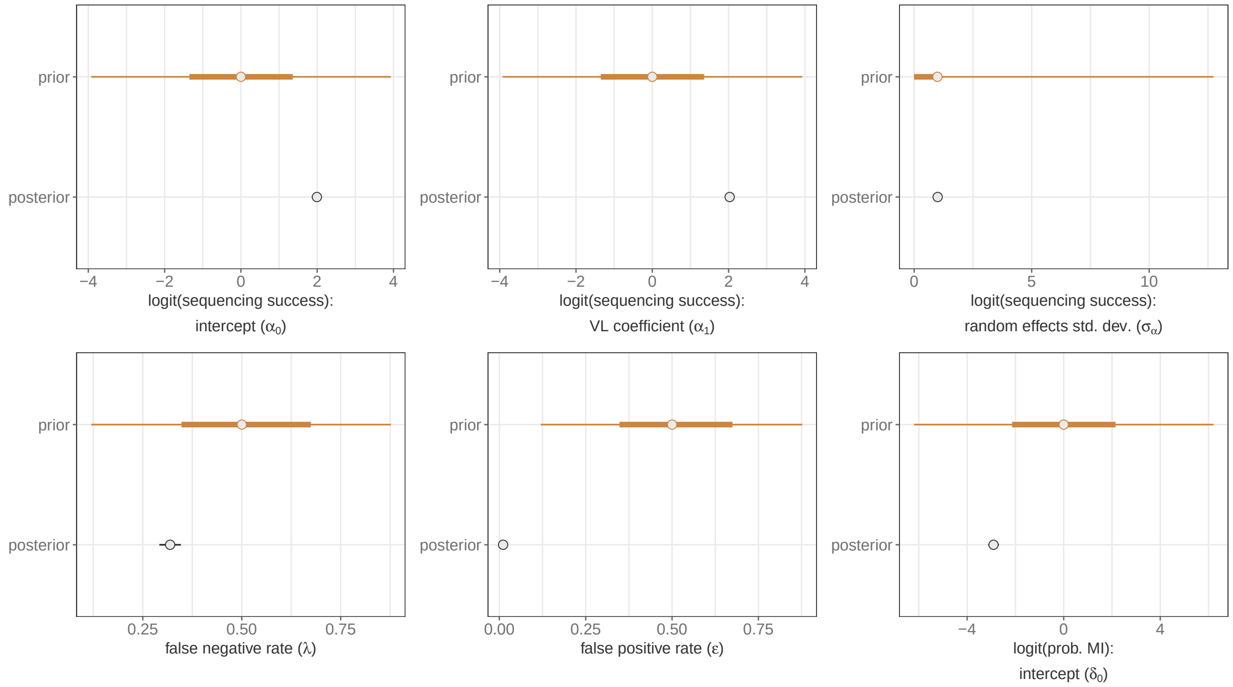


**Fig S.D. 9. Comparison of posterior and prior distributions of parameters in full model fit to simulated data with partial sequencing success and false positive or false negative multiple subgraph windows.** Median is plotted as the central estimate and horizontal bars extend to the 95% and 50% HPD. Some horizontal bars do not extend beyond the central point. Warm-up iterations are excluded. MI = multiple infection. VL = viral load ($\log_{10}$ copies/mL) standardized to mean = 0 and std. dev = 1. Std. dev. = standard deviation.

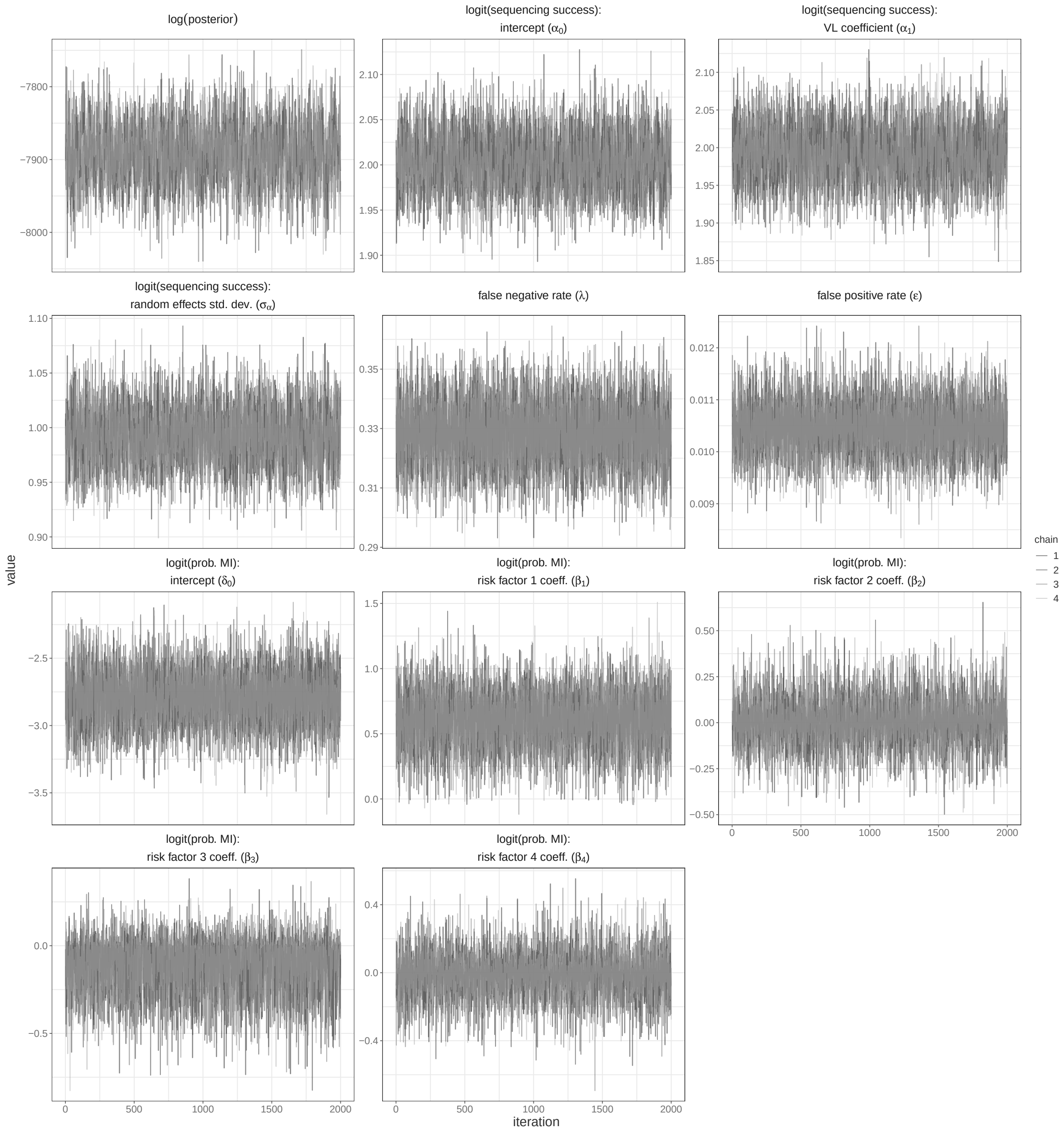


**Fig S.D. 10. MCMC trace plots for parameters in extended model fit to simulated data with partial sequencing success, false positive and false negative multiple subgraph windows, and a binary risk factor for harboring multiple infection.** Independent chains are shown in shades of grey. Warm-up iterations are excluded. MI = multiple infection. VL = viral load ($\log_{10}$ copies/mL) standardized to mean = 0 and std. dev = 1. Std. dev. = standard deviation.

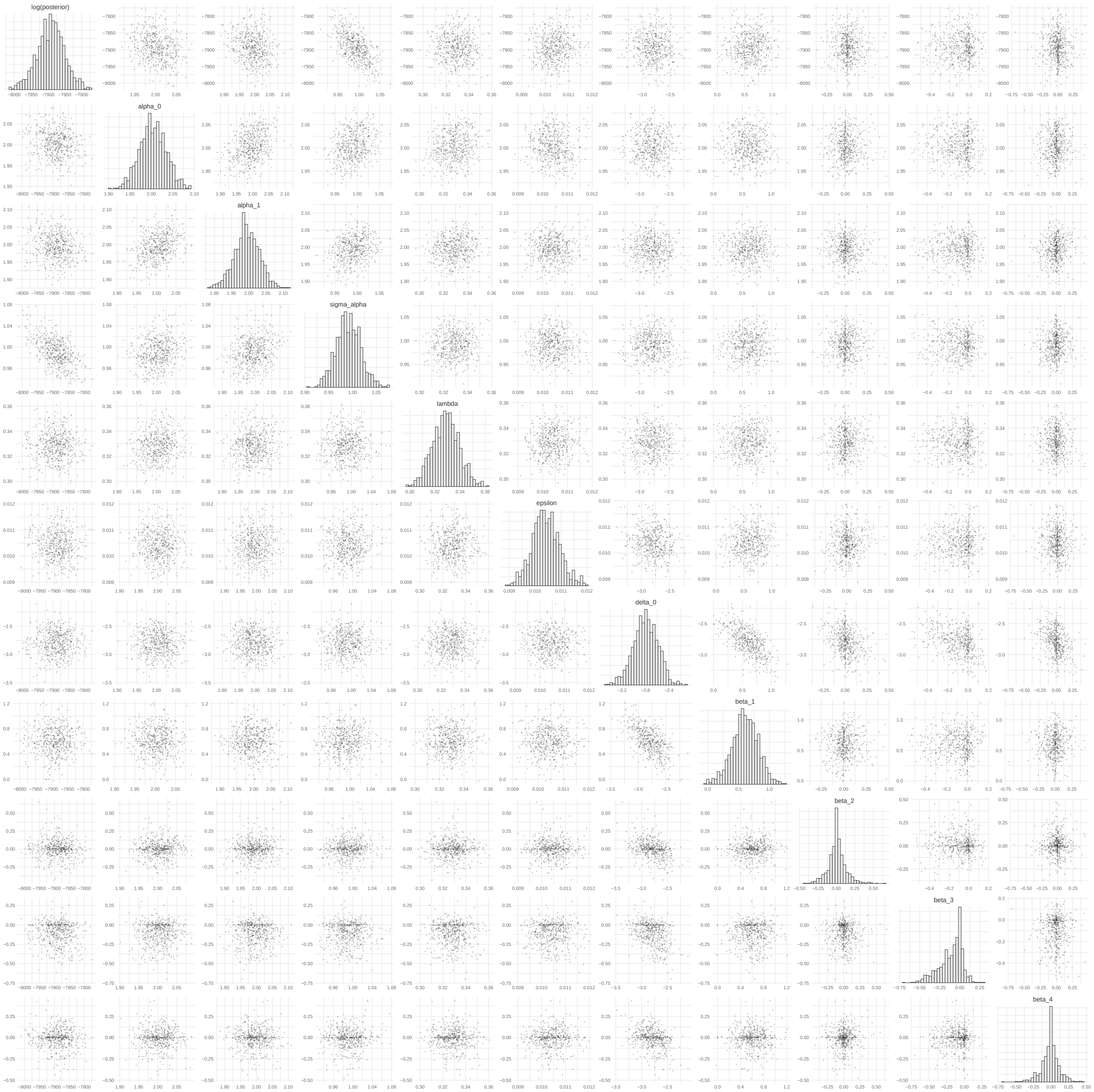


**Fig S.D. 11. MCMC pairs plots for parameters in extended model fit to simulated data with partial sequencing success, false positive and false negative multiple subgraph windows, and a binary risk factor for harboring multiple infection.** Independent chains are shown in shades of grey. Warm-up iterations are excluded. Includes a sample of 250 iterations per chain. MI = multiple infection.

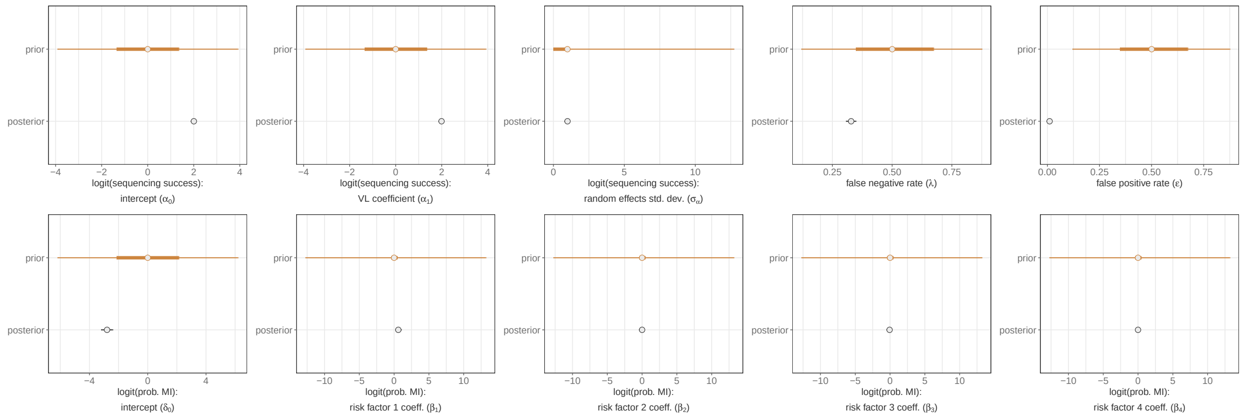


**Fig S.D. 12. Comparison of posterior and prior distributions of parameters in full model fit to simulated data with partial sequencing success, false positive or false negative multiple subgraph windows, and a binary risk factor for harboring multiple infection.** Median is plotted as the central estimate and horizontal bars extend to the 95% and 50% HPD. Some horizontal bars do not extend beyond the central point. Warm-up iterations are excluded. MI = multiple infection. VL = viral load ($\log_{10}$ copies/mL) standardized to mean = 0 and std. dev = 1. Std. dev. = standard deviation.

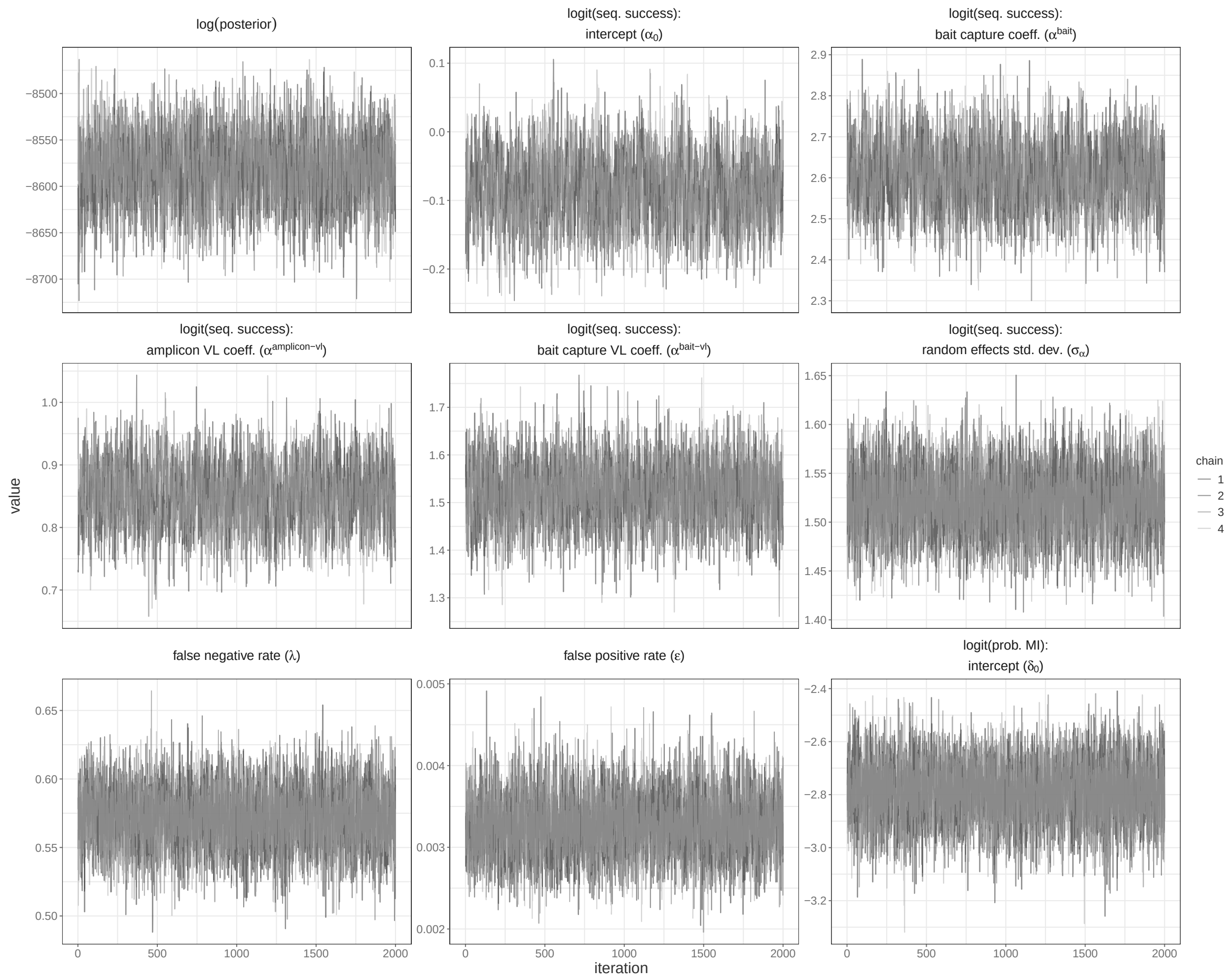


**Fig S.D. 13. MCMC trace plots for parameters in full model fit to deep sequence data generated from 2,029 Rakai Community Cohort Study participants living with viremic HIV.** Independent chains are shown in shades of grey. Warm-up iterations are excluded. MI = multiple infection. VL = viral load ($\log_{10}$ copies/mL) standardized to mean = 0 and std. dev = 1. Std. dev. = standard deviation.

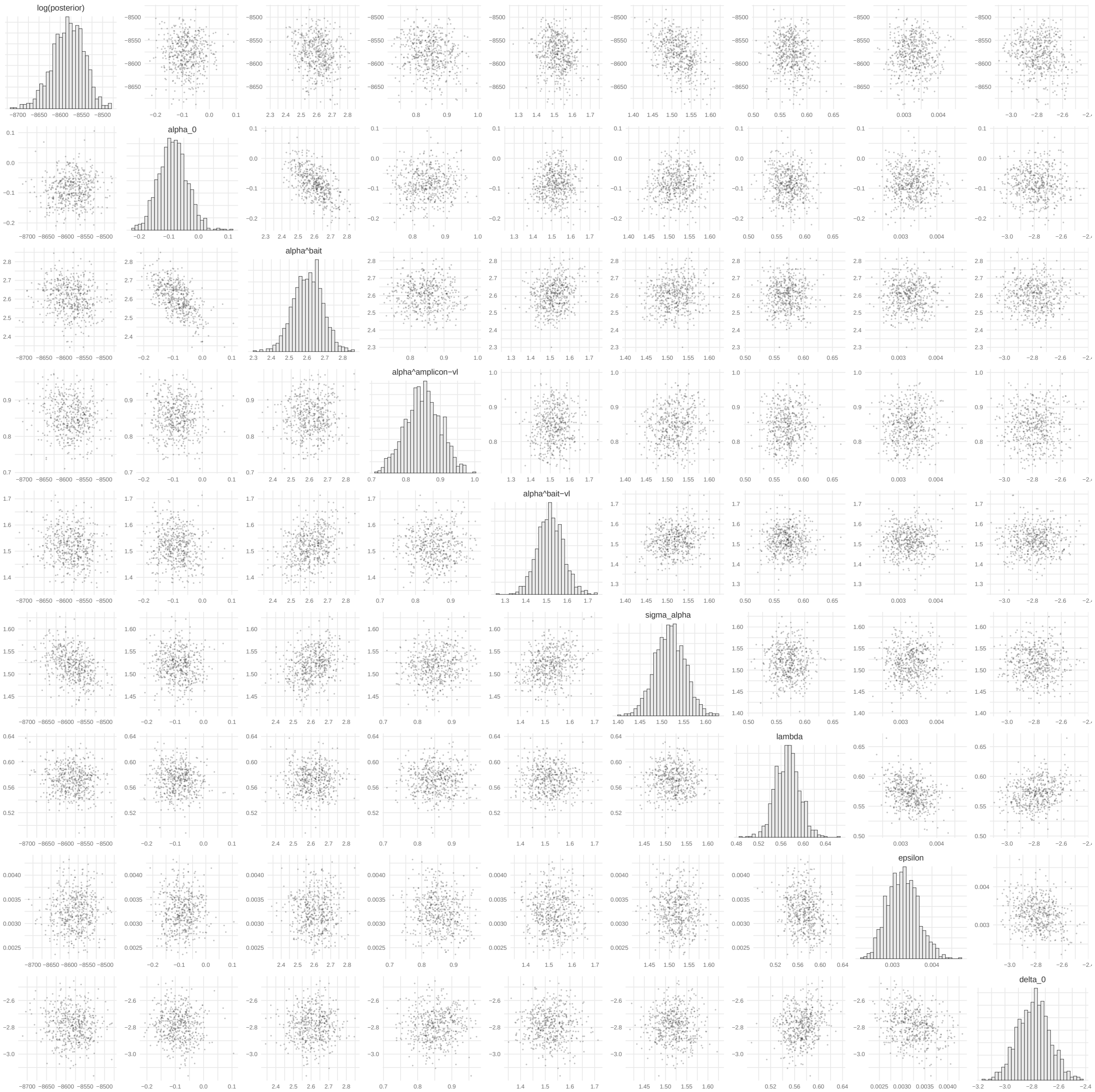


**Fig S.D. 14. MCMC pairs plots for parameters in full model fit to deep sequence data generated from 2,029 Rakai Community Cohort Study participants living with viremic HIV.** Independent chains are shown in shades of grey. Warm-up iterations are excluded. Includes a sample of 250 iterations per chain. MI = multiple infection.

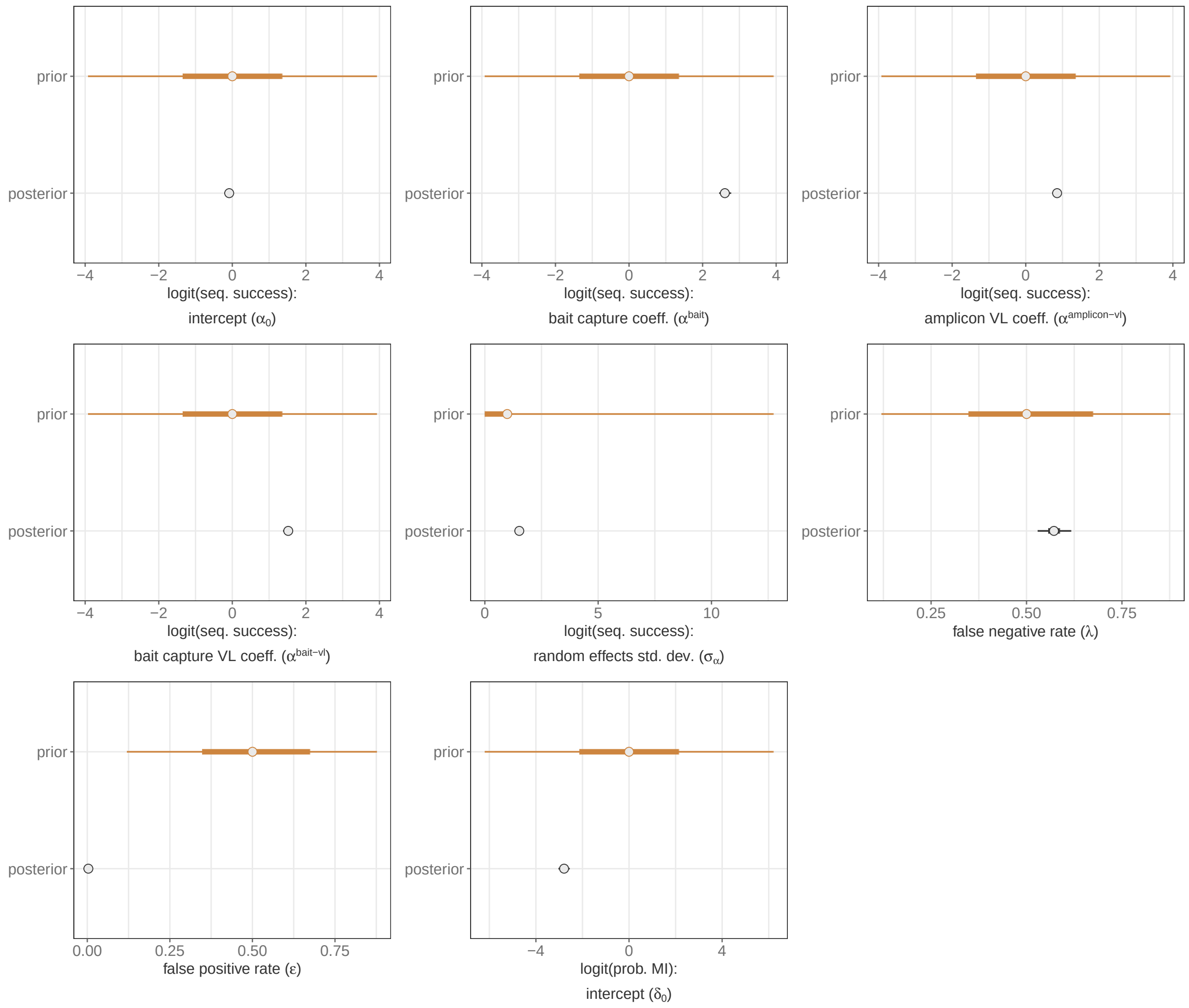


**Fig S.D. 15. Comparison of posterior and prior distributions of parameters in full model fit to deep sequence data generated from 2,029 Rakai Community Cohort Study participants living with viremic HIV.** Median is plotted as the central estimate and horizontal bars extend to the 95% and 50% HPD. Some horizontal bars do not extend beyond the central point. Warm-up iterations are excluded. MI = multiple infection. VL = viral load ($\log_{10}$ copies/mL) standardized to mean = 0 and std. dev = 1. Std. dev. = standard deviation.

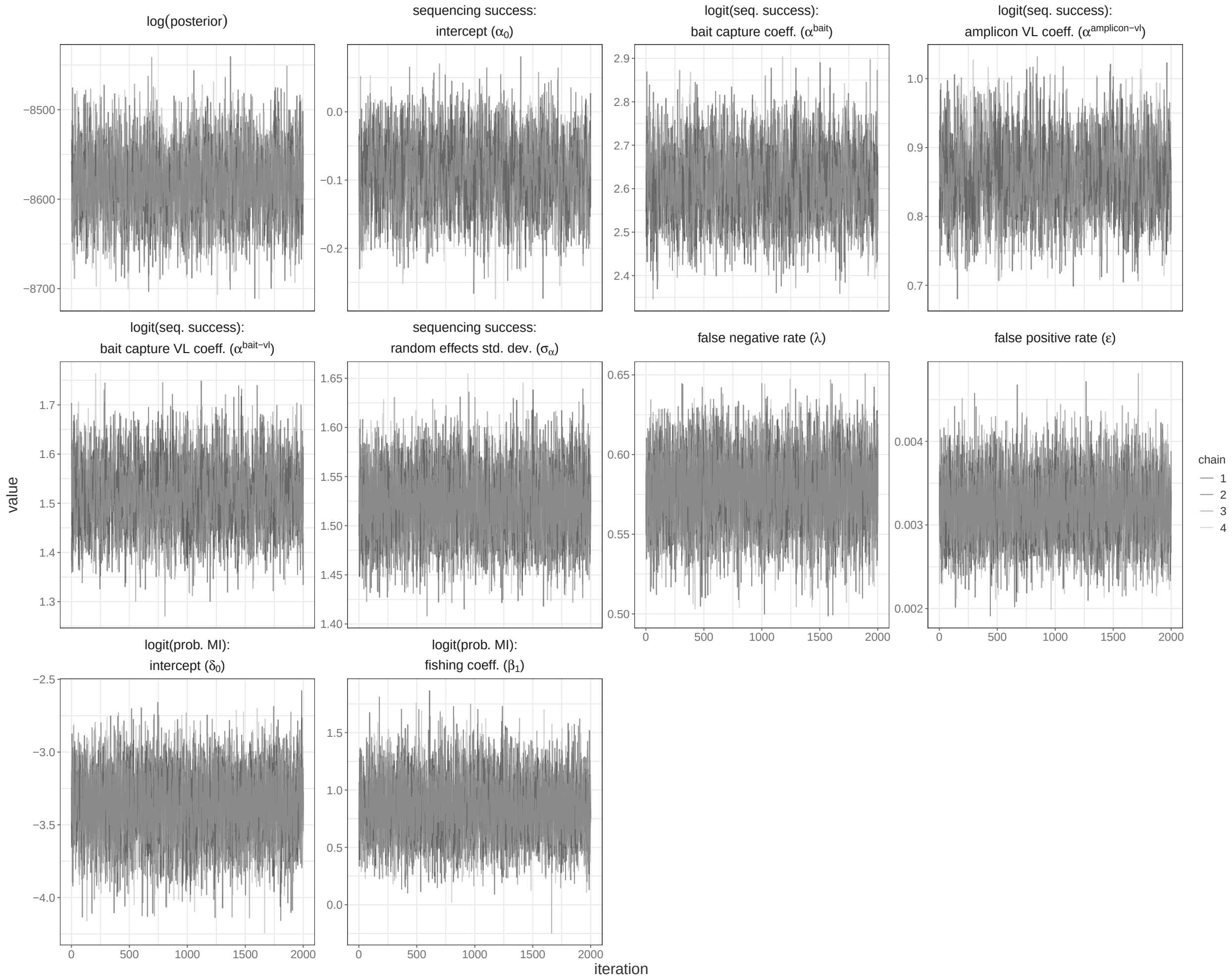


**Fig S.D. 16. MCMC trace plots for parameters in extended model with community type as a risk factor fit to deep sequence data generated from 2,029 Rakai Community Cohort Study participants living with viremic HIV.** Independent chains are shown in shades of grey. Warm-up iterations are excluded. MI = multiple infection. VL = viral load ($\log_{10}$ copies/mL) standardized to mean = 0 and std. dev = 1. Std. dev. = standard deviation.

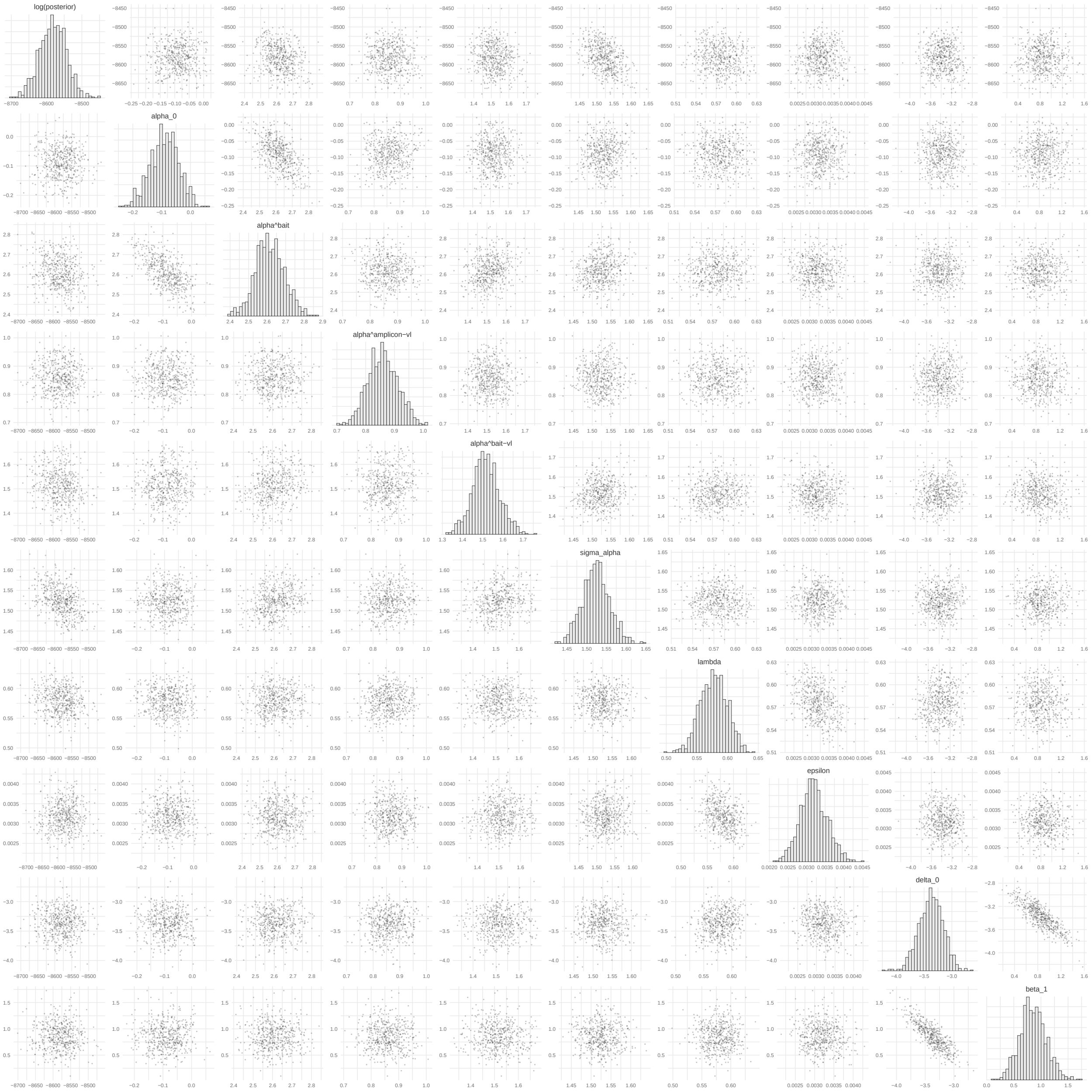


**Fig S.D. 17. MCMC pairs plots for parameters in extended model with community type as a risk factor fit to deep sequence data generated from 2,029 Rakai Community Cohort Study participants living with viremic HIV.** Independent chains are shown in shades of grey. Warm-up iterations are excluded. Includes a sample of 250 iterations per chain. MI = multiple infection.

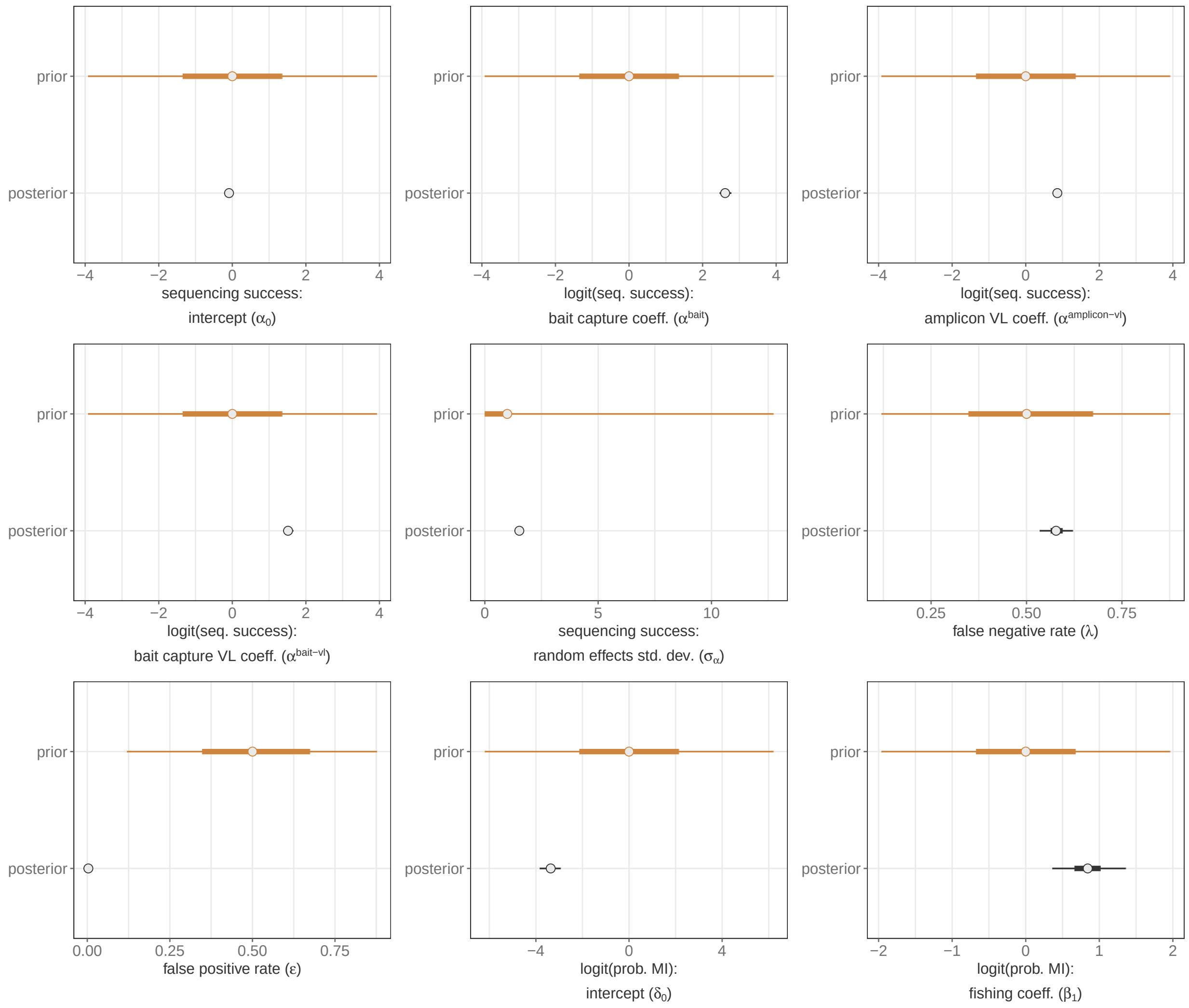


**Fig S.D. 18. Comparison of posterior and prior distributions of parameters in extended model with community type as a risk factor fit to deep sequence data generated from 2,029 Rakai Community Cohort Study participants living with viremic HIV.** Median is plotted as the central estimate and horizontal bars extend to the 95% and 50% HPD. Some horizontal bars do not extend beyond the central point. Warm-up iterations are excluded. MI = multiple infection. VL = viral load ($\log_{10}$ copies/mL) standardized to mean = 0 and std. dev = 1. Std. dev. = standard deviation.

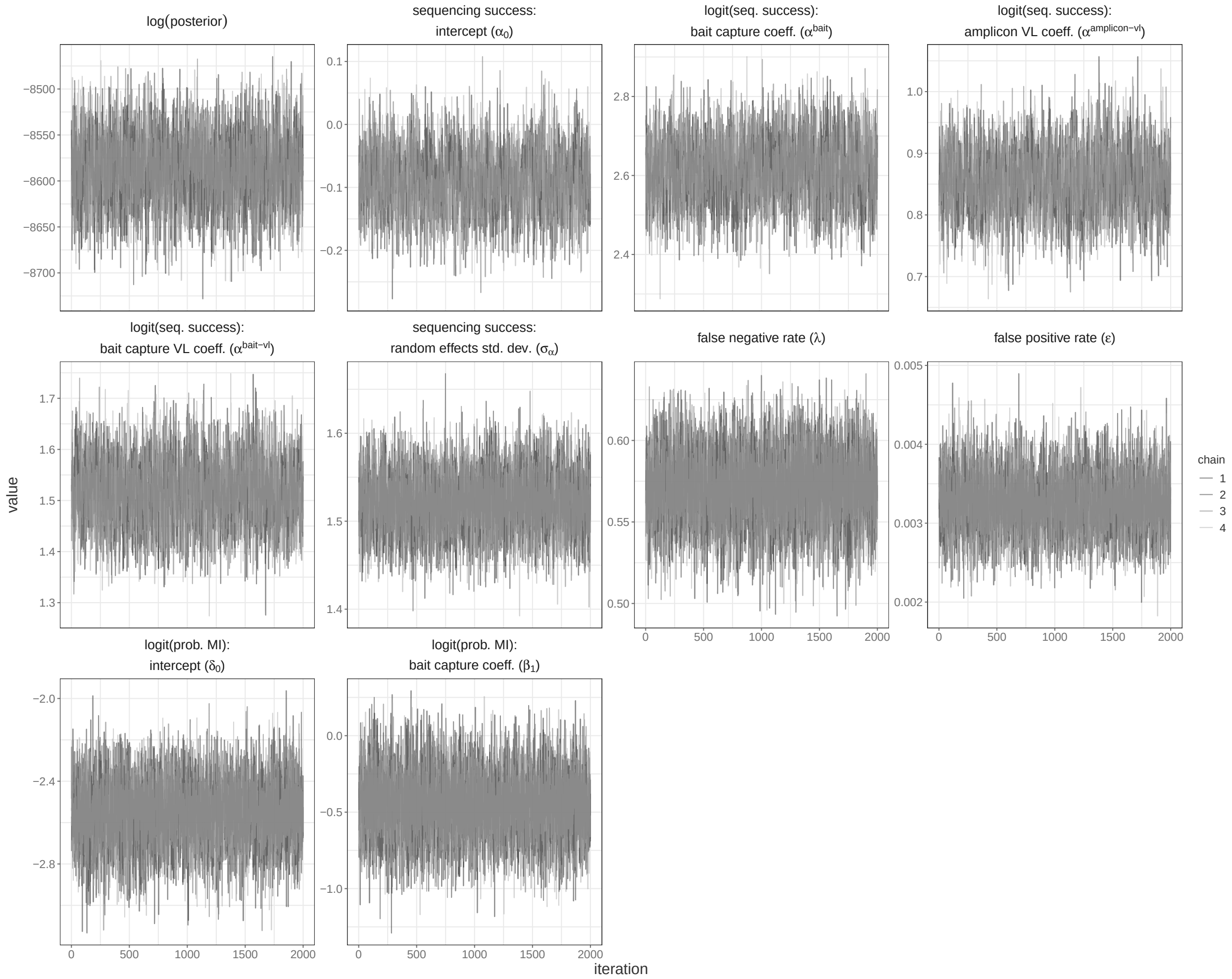


**Fig S.D. 19. MCMC trace plots for parameters in extended model with deep-sequencing protocol as a risk factor fit to deep sequence data generated from 2,029 Rakai Community Cohort Study participants living with viremic HIV.** Independent chains are shown in shades of grey. Warm-up iterations are excluded. MI = multiple infection. VL = viral load ($\log_{10}$ copies/mL) standardized to mean = 0 and std. dev = 1. Std. dev. = standard deviation.

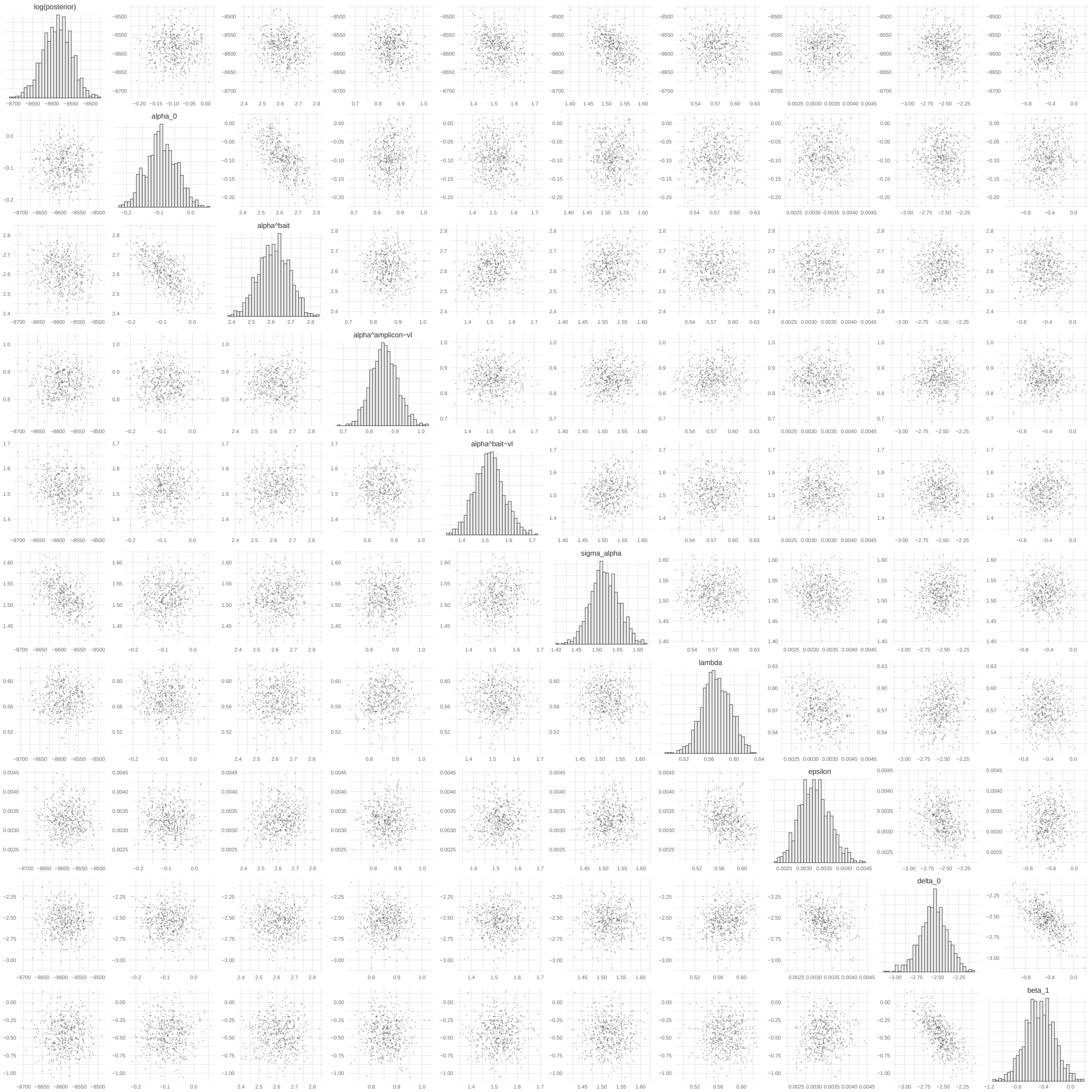


**Fig S.D. 20. MCMC pairs plots for parameters in extended model withdeep-sequencing protocol as a risk factor fit to deep sequence data generated from 2,029 Rakai Community Cohort Study participants living with viremic HIV.** Independent chains are shown in shades of grey. Warm-up iterations are excluded. Includes a sample of 250 iterations per chain. MI = multiple infection.

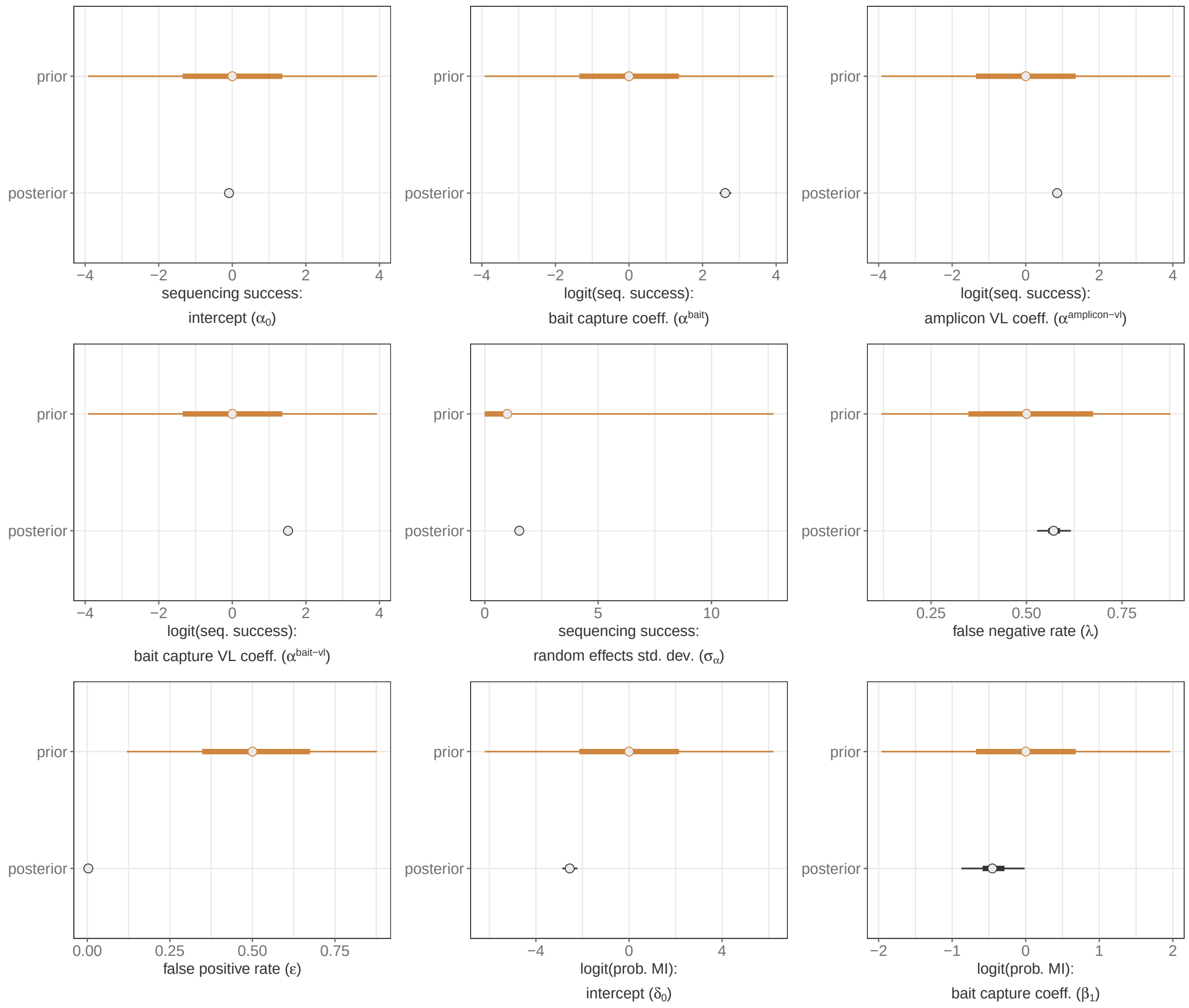


**Fig S.D. 21. Comparison of posterior and prior distributions of parameters in extended model with sequencing technology as a risk factor fit to deep sequence data generated from 2,029 Rakai Community Cohort Study participants living with viremic HIV.** Median is plotted as the central estimate and horizontal bars extend to the 95% and 50% HPD. Some horizontal bars do not extend beyond the central point. Warm-up iterations are excluded. MI = multiple infection. VL = viral load ($\log_{10}$ copies/mL) standardized to mean = 0 and std. dev = 1. Std. dev. = standard deviation.

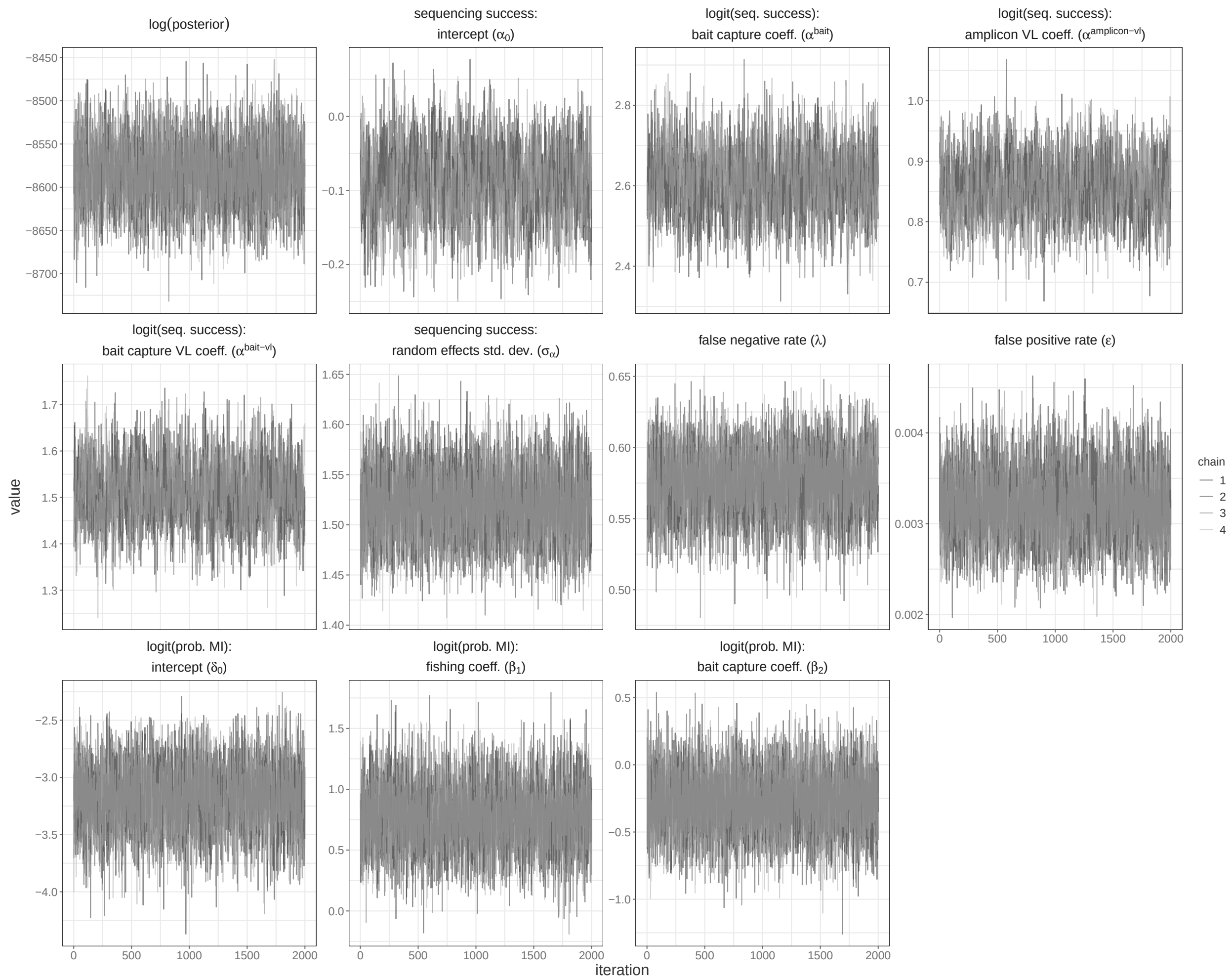


**Fig S.D. 22. MCMC trace plots for parameters in extended model with deep-sequencing protocol and community type as a risk factor fit to deep sequence data generated from 2,029 Rakai Community Cohort Study participants living with viremic HIV.** Independent chains are shown in shades of grey. Warm-up iterations are excluded. MI = multiple infection. VL = viral load ($\log_{10}$ copies/mL) standardized to mean = 0 and std. dev = 1. Std. dev. = standard deviation.

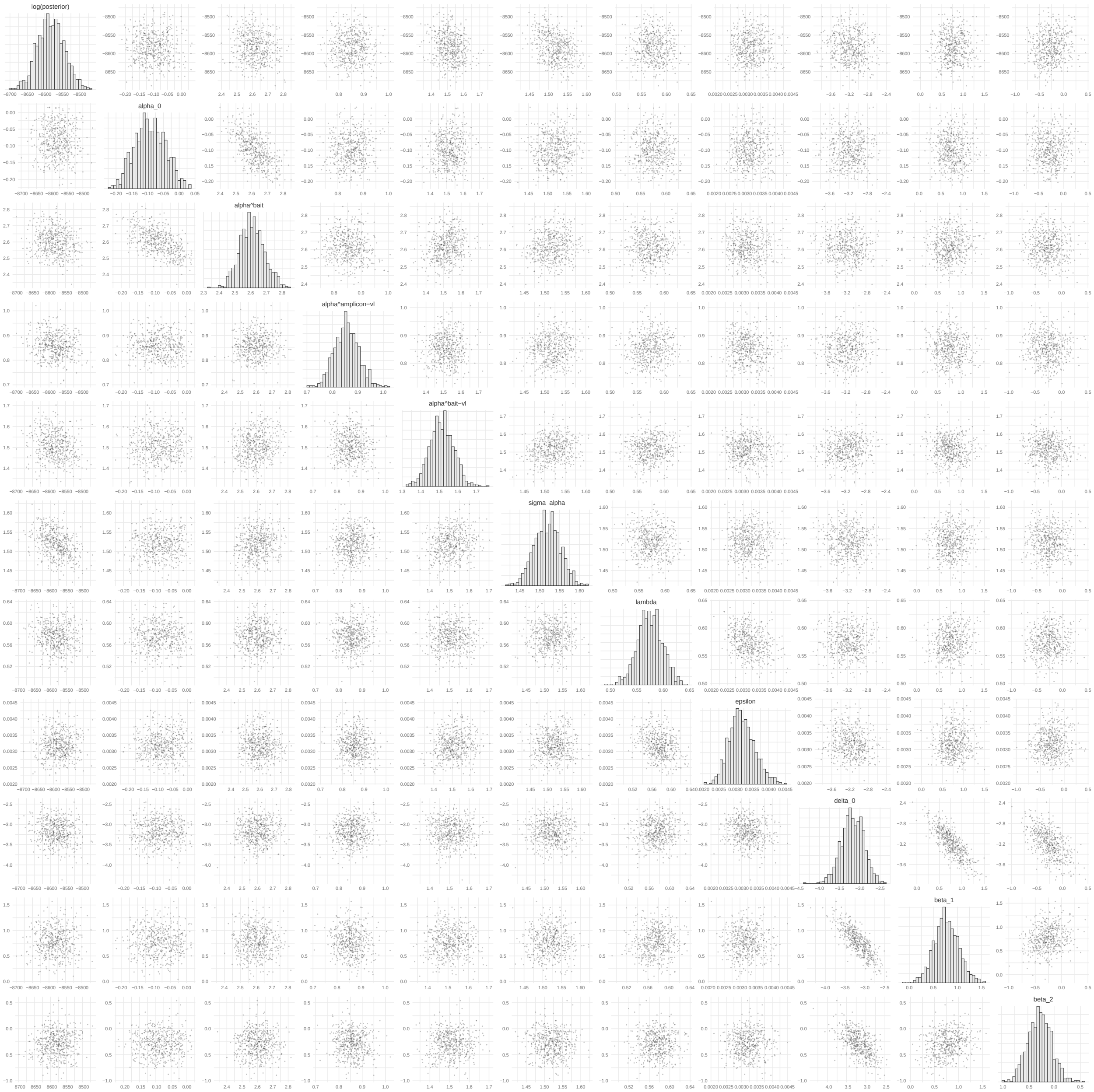


**Fig S.D. 23. MCMC pairs plots for parameters in extended model withdeep-sequencing protocol and community type as a risk factor fit to deep sequence data generated from 2,029 Rakai Community Cohort Study participants living with viremic HIV.** Independent chains are shown in shades of grey. Warm-up iterations are excluded. Includes a sample of 250 iterations per chain. MI = multiple infection.

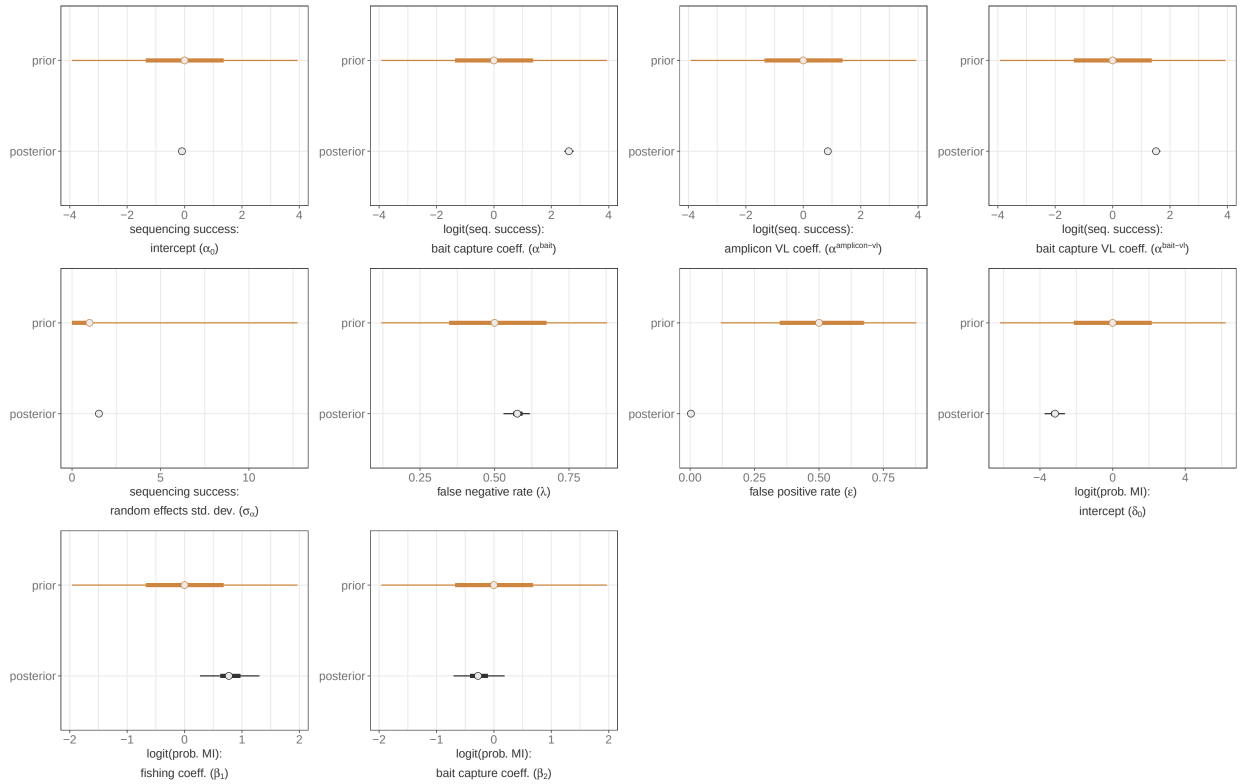


**Fig S.D. 24. Comparison of posterior and prior distributions of parameters in extended model with sequencing technology and community type as a risk factor fit to deep sequence data generated from 2,029 Rakai Community Cohort Study participants living with viremic HIV.** Median is plotted as the central estimate and horizontal bars extend to the 95% and 50% HPD. Some horizontal bars do not extend beyond the central point. Warm-up iterations are excluded. MI = multiple infection. VL = viral load ($\log_{10}$ copies/mL) standardized to mean = 0 and std. dev = 1. Std. dev. = standard deviation.

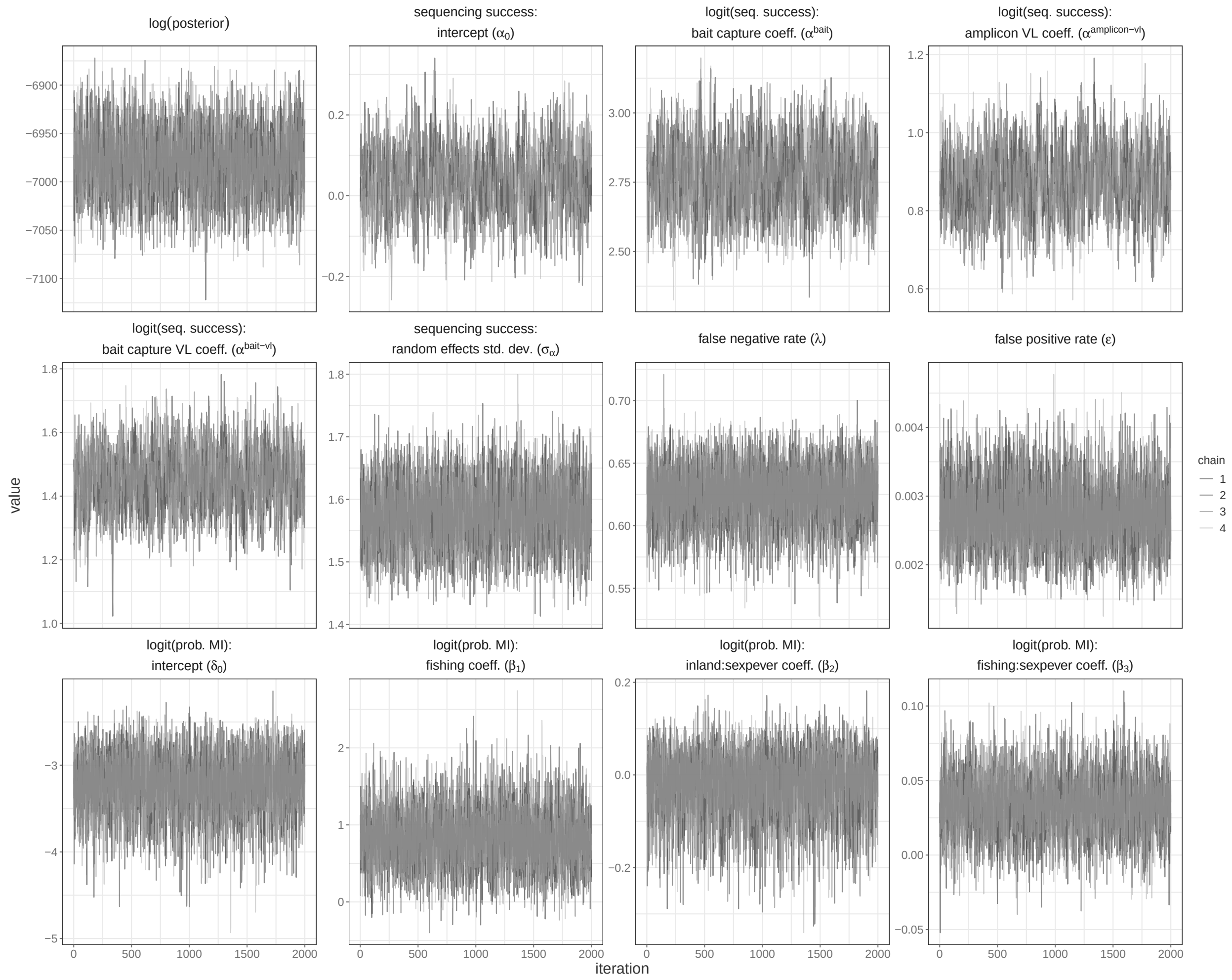


**Fig S.D. 25. MCMC trace plots for parameters in extended model with community type and lifetime sex partners as a risk factor for harboring multiple infections fit to deep sequence data generated from 997 men living with viremic HIV who participated in the Rakai Community Cohort Study.** Independent chains are shown in shades of grey. Warm-up iterations are excluded. MI = multiple infection. VL = viral load ($\log_{10}$ copies/mL) standardized to mean = 0 and std. dev = 1. Std. dev. = standard deviation.

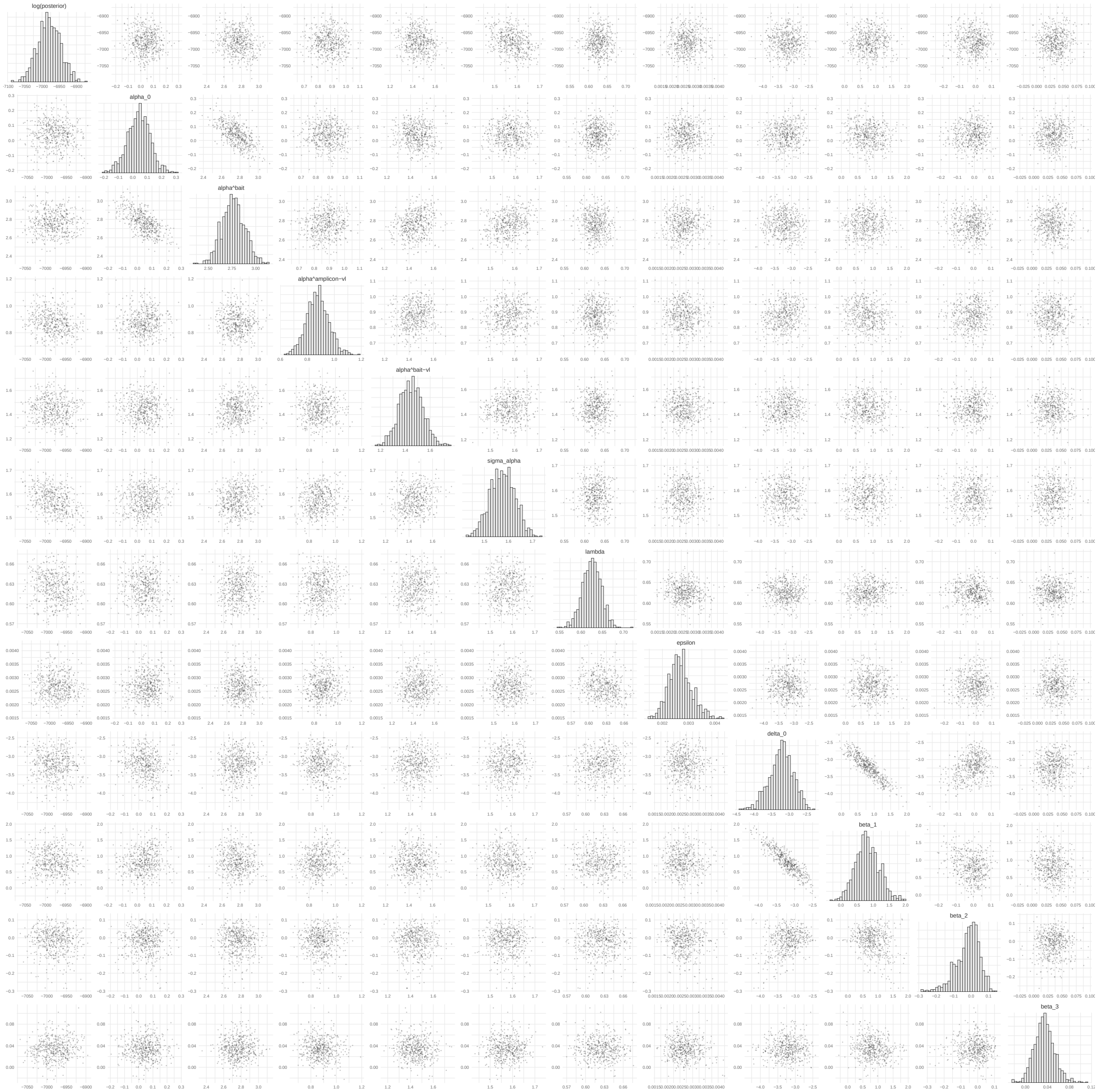


**Fig S.D. 26. MCMC pairs plots for parameters in extended model with community type and lifetime sex partners as a risk factor for harboring multiple infections fit to deep sequence data generated from 997 men living with viremic HIV who participated in the Rakai Community Cohort Study.** Independent chains are shown in shades of grey. Warm-up iterations are excluded. Includes a sample of 250 iterations per chain. MI = multiple infection.

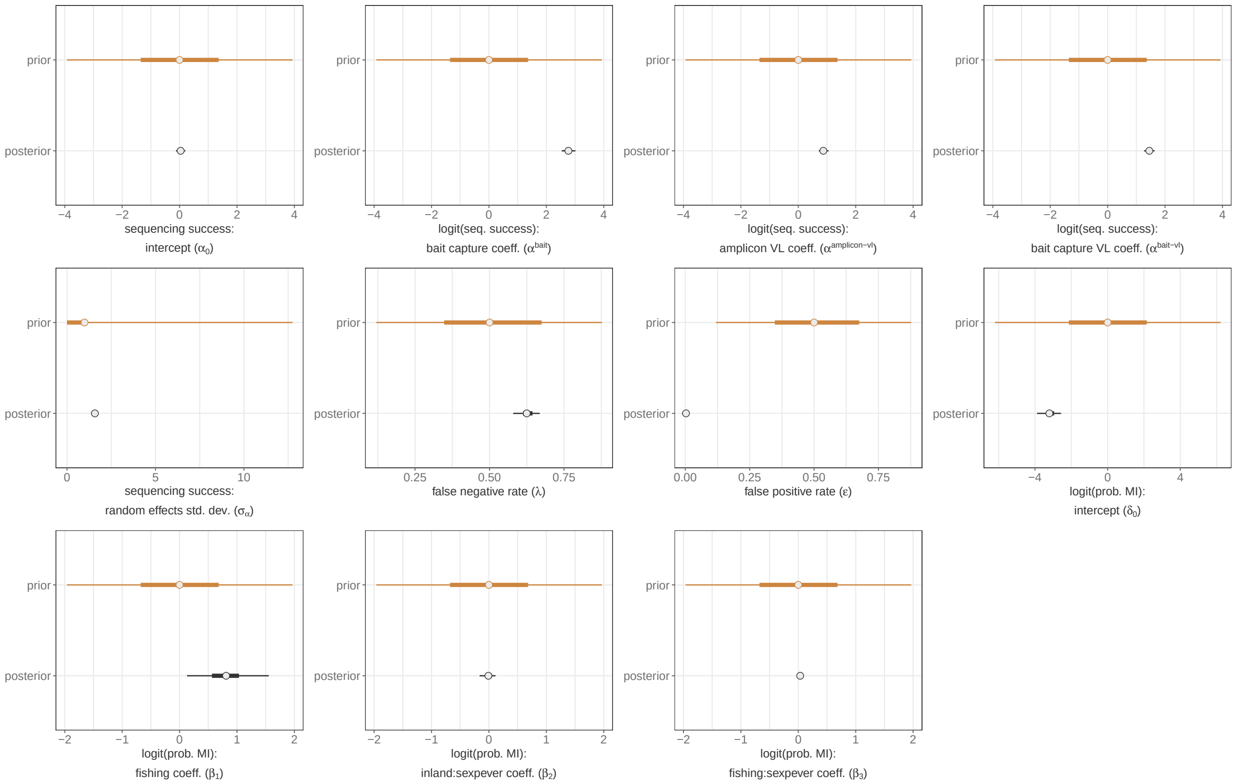


**Fig S.D. 27. Comparison of posterior and prior distributions of parameters in extended model with sequencing technology as a risk factor fit to deep sequence data generated from 997 Rakai Community Cohort Study participants living with viremic HIV.** Median is plotted as the central estimate and horizontal bars extend to the 95% and 50% HPD. Some horizontal bars do not extend beyond the central point. Warm-up iterations are excluded. MI = multiple infection. VL = viral load ($\log_{10}$ copies/mL) standardized to mean = 0 and std. dev = 1. Std. dev. = standard deviation.

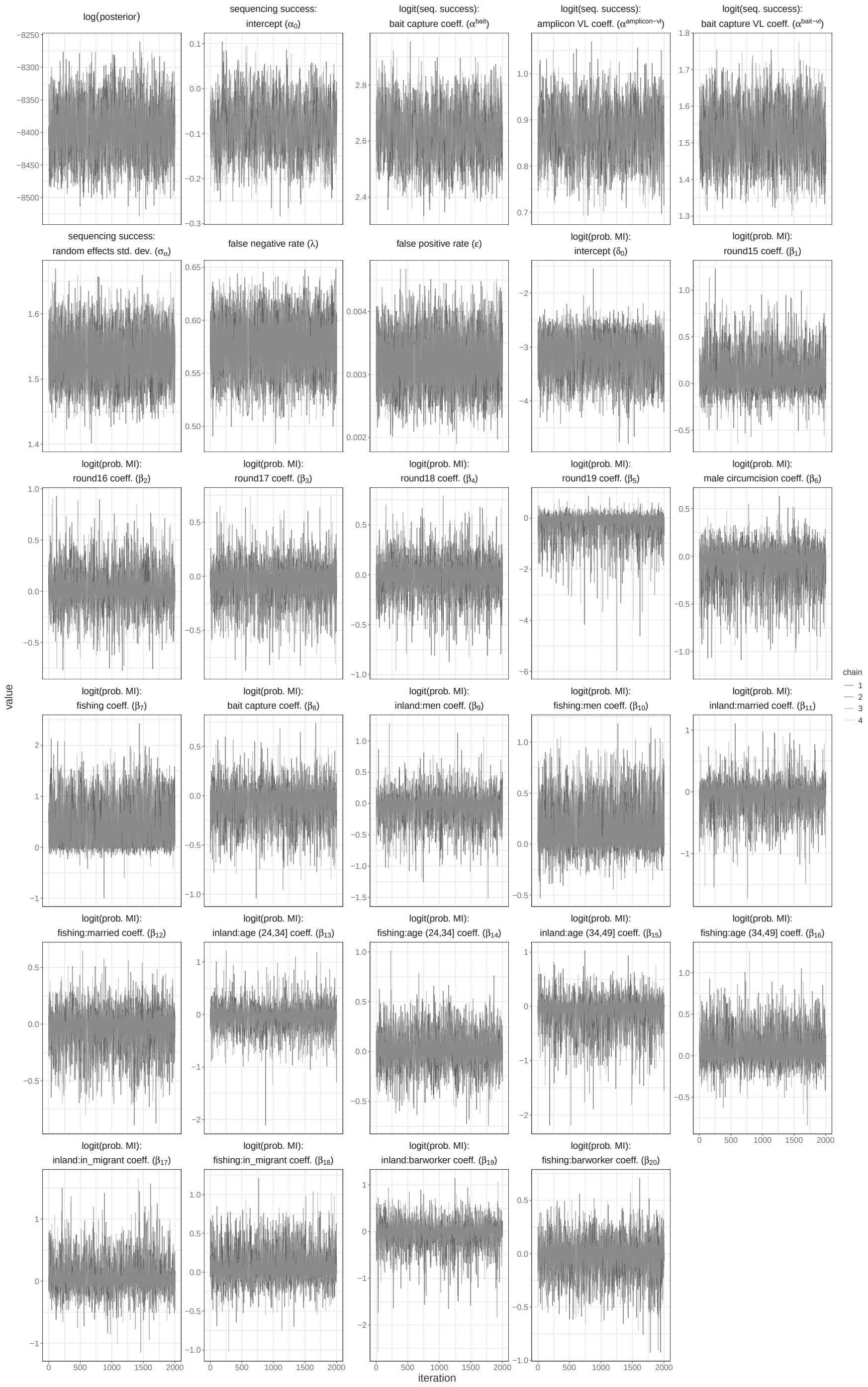

**Fig S.D. 28. MCMC trace plots for parameters in extended model with variable selection to identify risk factor for harboring multiple infections fit to deep sequence data generated from 1,970 Rakai Community Cohort Study participants living with viremic HIV.** Independent chains are shown in shades of grey. Warm-up iterations are excluded. MI = multiple infection. VL = viral load ($\log_{10}$ copies/mL) standardized to mean = 0 and std. dev = 1. Std. dev. = standard deviation.

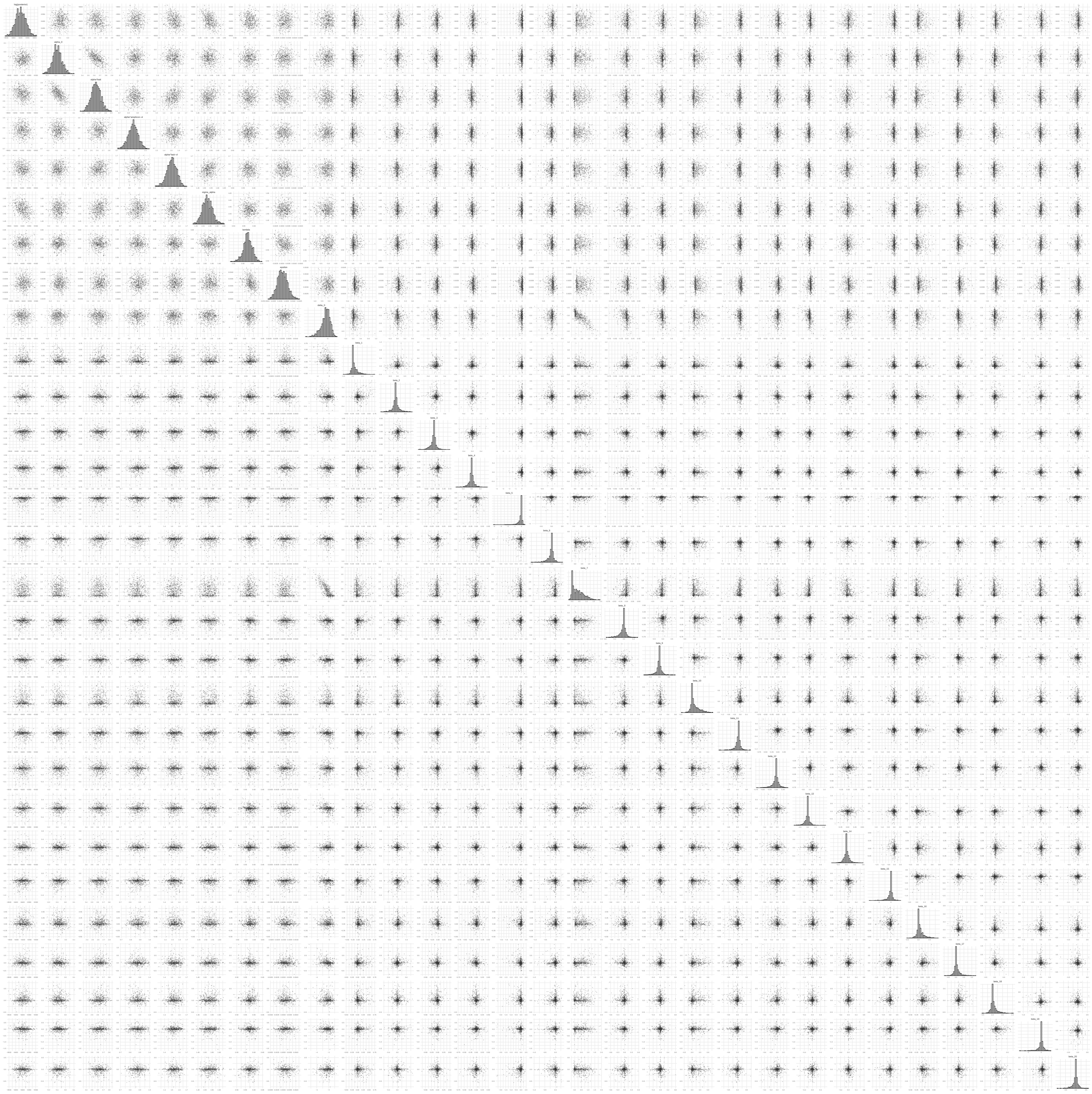

**Fig S.D. 29. MCMC pairs plots for parameters in extended model with variable selection to identify risk factor for harboring multiple infections fit to deep sequence data generated from 1,970 Rakai Community Cohort Study participants living with viremic HIV.** Independent chains are shown in shades of grey. Warm-up iterations are excluded. Includes a sample of 250 iterations per chain. MI = multiple infection.

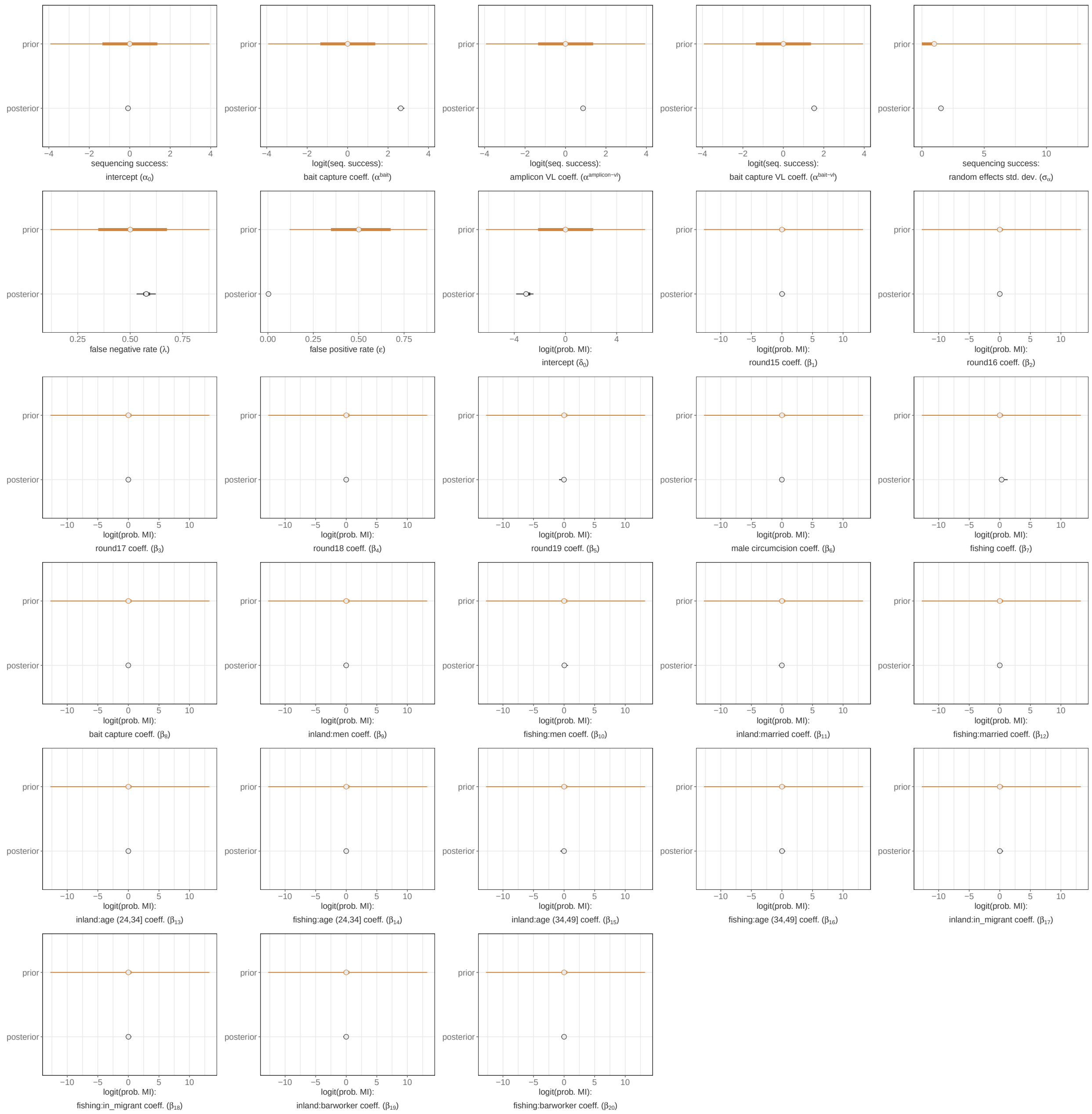


**Fig S.D. 30. Comparison of posterior and prior distributions of parameters in extended model with variable selection to identify risk factor fit to deep sequence data generated from 1,970 Rakai Community Cohort Study participants living with viremic HIV.** Median is plotted as the central estimate and horizontal bars extend to the 95% and 50% HPD. Some horizontal bars do not extend beyond the central point. Warm-up iterations are excluded. MI = multiple infection. VL = viral load ($\log_{10}$ copies/mL) standardized to mean = 0 and std. dev = 1. Std. dev. = standard deviation.
