## Supplementary material for "Quantifying prevalence and risk factors of HIV multiple infection in Uganda from population-based deep-sequence data": S5 File

#### 1 Choice of genome window

Phylogenetic trees from the HIV deep-sequence reads generated from 2,029 Rakai Community Cohort Study participants living with viremic HIV using *phyloscanner*. In total, phylogenetic trees were inferred using data from 287 250 base pair (bp) overlapping windows with a 25 bp step between windows. To generate a set of independent data points from each sequenced participant-visit we first downsampled the 287 overlapping windows to a set of 29 250 bp windows beginning at position 800 in the HXB2 (GenBank: K03455.1) genome. Summarized data in the form of the genome coverage in the phyloscanner output ($N_i^{obs}$) and the number of windows with multiple phylogenetic subgraphs ($MI_i^{obs}$) was highly correlated between the set of full windows and the set of non-overlapping windows (Pearson's $\rho = 1$, 0.97, respectively, Genome Window Fig S.G.1).

#### 2 Alternate set of non-overlapping windows

Based on the window size and window step size, there are 10 potential sets of non-overlapping windows that could have been chosen to generate a set of independent data-points for each sequenced participant visits. To assess the sensitivity of our results to the choice of non-overlapping windows we alternatively downsampled the data to the set of non-overlapping 250 bp windows beginning at position 950 in the HXB2 genome.

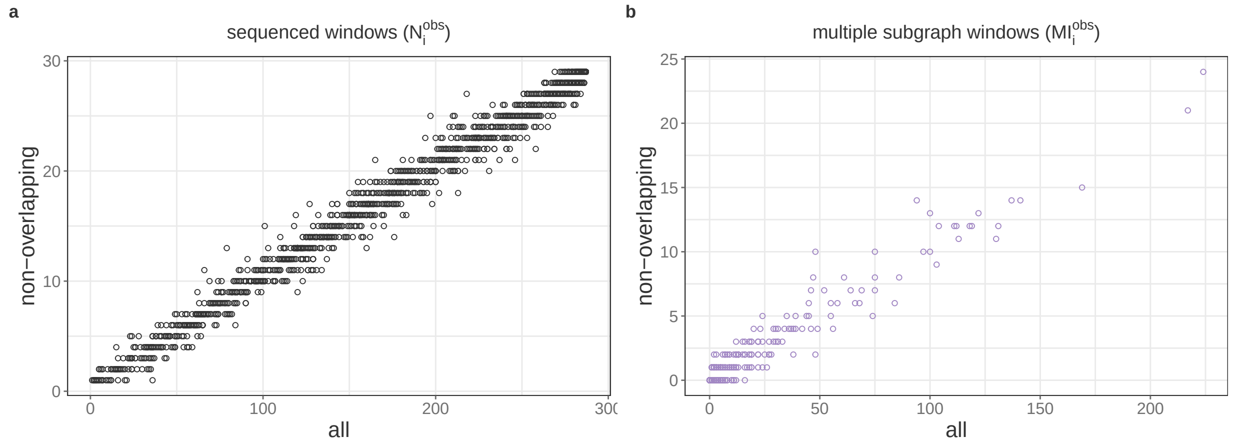


**Fig S.G.1. Correlation between summary statistics from within-host deep sequence phylogenies from all genome windows and non-overlapping windows generated from 2,029 Rakai Community Cohort Study living with viremic HIV.** (A) Number of windows with phyloscanner output for each participant-visit. (B) Number of windows with multiple phylogenetic subgraphs for each participant-visit.

Genome coverage and the number of windows with multiple phylogenetic subgraphs was highly correlated between the two sets of non-overlapping windows (Pearson's $\rho = 0.99$, 0.94, respectively).

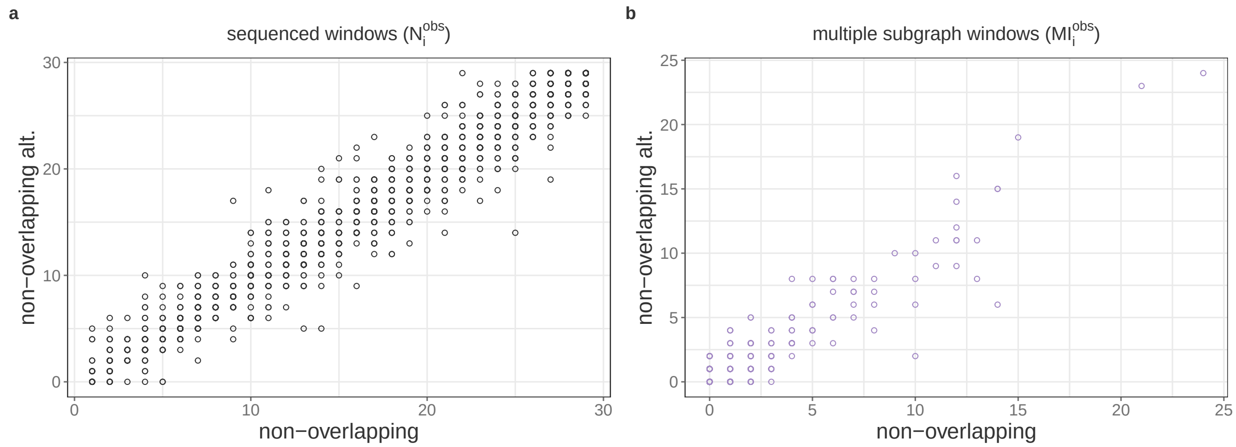


**Fig S.G.2. Correlation between summary statistics from within-host deep sequence phylogenies from all genome windows and non-overlapping windows generated from 2,029 Rakai Community Cohort Study participant-visits living with viremic HIV.** (A) Number of windows with phyloscanner output for each participant-visit. (B) Number of windows with multiple phylogenetic subgraphs for each participant-visit.

### 3 Sensitivity of results to non-overlapping windows

Finally, we fit our inference model accounting for partial sequencing success, false negative multiple subgraph windows, and false positive multiple subgraph windows to data from the two sets of non-overlapping genome windows (Table S.G.1-S.G.2). We note that parameter estimates from the two model fits are highly similar, indicating that our results are not particularly sensitive to the exact choice of genome window sets.

| Parameter | Prior | Median (95% HPD) | Bulk ESS | Tail ESS | $\hat{R}$ |
|---|---|---|---|---|---|
| $\alpha_0$ | Normal(0,$2^2$) | -0.08 (-0.18, 0.01) | 1094.34 | 2116.52 | 1 |
| $\alpha^{bait}$ | Normal(0,$2^2$) | 2.61 (2.45, 2.76) | 1522.61 | 3106.93 | 1 |
| $\alpha^{amplicon-vl}$ | Normal(0,$2^2$) | 0.85 (0.76, 0.95) | 1513.14 | 2991.73 | 1 |
| $\alpha^{bait-vl}$ | Normal(0,$2^2$) | 0.85 (0.76, 0.95) | 1513.14 | 2991.73 | 1 |
| $\sigma_\alpha$ | Half-Cauchy(0,1) | 1.52 (1.45, 1.59) | 4631.32 | 6140 | 1 |
| $\delta_0$ | Normal(0,$3.16^2$) | -2.79 (-3.03, -2.56) | 7236.45 | 4682.27 | 1 |
| logit($\lambda$) | Normal(0,1) | 0.29 (0.12, 0.47) | 5004.3 | 5833.33 | 1 |
| logit($\epsilon$) | Normal(0,1) | -5.73 (-5.96, -5.5) | 4917.44 | 5128.91 | 1 |

**Table S.G.1. Parameter estimates for full model fit to deep sequence data from 2,029 RCCS participants living with viremic HIV.** ESS = effective sample size. HPD = highest posterior density.

| Parameter | Prior | Median (95% HPD) | Bulk ESS | Tail ESS | $\hat{R}$ |
|---|---|---|---|---|---|
| $\alpha_0$ | Normal(0,$2^2$) | -0.14 (-0.24, -0.05) | 1543.99 | 2884.32 | 1 |
| $\alpha^{bait}$ | Normal(0,$2^2$) | 2.68 (2.52, 2.84) | 1949.32 | 3238.59 | 1 |
| $\alpha^{amplicon-vl}$ | Normal(0,$2^2$) | 0.84 (0.74, 0.94) | 1391.82 | 2530.72 | 1 |
| $\alpha^{bait-vl}$ | Normal(0,$2^2$) | 1.52 (1.4, 1.65) | 2138.08 | 3570.24 | 1 |
| $\sigma_\alpha$ | Half-Cauchy(0,1) | 1.5 (1.44, 1.57) | 4984.07 | 5920.38 | 1 |
| $\delta_0$ | Normal(0,$3.16^2$) | -2.76 (-3, -2.54) | 7571.4 | 5842.81 | 1 |
| logit($\lambda$) | Normal(0,1) | 0.48 (0.32, 0.64) | 6901.83 | 6604.32 | 1 |
| logit($\epsilon$) | Normal(0,1) | -5.83 (-6.08, -5.58) | 4772.54 | 6006.13 | 1 |

**Table S.G.2. Parameter estimates for full model fit to deep sequence data from 2,029 RCCS participants living with viremic HIV from alternative non-overlapping genome windows.** ESS = effective sample size. HPD = highest posterior density.
